# Molecular Profiling of a Triple-negative Breast Cancer Cohort in India for EGFR and AR expression analyzed for patient outcomes showed a distinct subset of cellular co-expression

**DOI:** 10.1101/2024.10.25.24316141

**Authors:** Pooja Vaid, Anirudha Puntambekar, Pranali Kanse, Shweta Kadu, Piyush Agrawal, Aditi Khatpe, Rituja Banale, Ruhi Reddy, Devaki A. Kelkar, Sridhar Hannenhalli, L S Shashidhara, Chaitanyanand Koppiker, Madhura Kulkarni

**Affiliations:** Centre for Translational Cancer Research: a joint initiative of Indian Institute of Science Education and Research (IISER) Pune and Prashanti Cancer Care Mission (PCCM) Pune; Department of Biological Sciences, Ashoka University, Sonipat; KRSNAA Diagnostics Ltd., Pune; Prashanti Cancer Care Mission, Pune; Department of Medical Research, SRM Medical College Hospital & Research Centre, SRMIST, Kattankulathur, Chennai; School of Medicine, IUPUI, Indiana, USA; Cancer Data Science Laboratory, Center for Cancer Research, NCI, NIH, Bethesda; Department of Biology, Indian Institute of Science Education and Research, Pune; National Centre for Biological Sciences, Bangalore

**Author notes:** Corresponding author: Madhura Kulkarni, PhD, Senior Scientist and DBT-Ramalingaswami Fellow, Centre for Translational Cancer Research., Indian Institute of Science Education and Research (IISER) Pune and Prashanti Cancer Care Mission, Pune, B-403, Third Floor, Main Building, Dr Homi Bhabha Road, Pashan, Pune 411 008 Maharashtra INDIA.

**Keywords:** Basal TNBC, EGFR, AR, co-expression of EGFR and AR

## Abstract

TNBC is the most aggressive breast cancer subtype, with a higher recurrence rate. TNBC proportion within breast cancer incidence in the Indian population is 22-30%. Despite the high incidence rate, the heterogeneity within TNBC subtype in Indian cohorts is not studied at scale.

Here, leveraging an Indian cohort of 93 TNBC patients, we evaluated the basal and LAR subtypes in terms of the expression of known markers such as EGFR and AR and further assessed the association of marker gene expression with patient outcome and treatment response.

In our cohort, 65% of the patients were EGFR-positive, 38% had positive AR expression, where both the subsets showed shorter disease-free survival outcomes. Additionally, 25% of the cohort showed AR and EGFR co-expression. Upon closer observation, using IHC and duplex staining, we noted that 15% of the tumors, in fact, had double-positive cancer cells, i.e., cellular co-expression of AR and EGFR. Patients with double-positive cells had poorer disease-free survival compared to the ones with the tissue-level co-expression of EGFR and AR but without cellular co-expression. The presence of EGFR+AR+ double-positive cells was further validated in publicly available single-cell data sets for TNBC patients from other ethnic backgrounds, albeit to a lesser extent than what was observed in our Indian cohort. Overall, our results highlight the heterogeneous nature of Indian TNBC tumors and provide further insight into ethnic variation in TNBC presentation that can be further exploited for precision and personalized targeted therapy.

---

Triple-negative breast cancer (TNBC) is a clinically aggressive subtype of breast cancer, i.e., it presents more often with high-grade disease, node involvement, and up to 42% of the patients recur within two years of primary diagnosis ^1–3^. TNBC in India, due to the lack of targetable markers and unreasonable costs for newly introduced immunotherapy, is still largely treated with standard cytotoxic chemotherapy regimens with only 25-35% percent responding to treatment (Das et al, 2024, ^4^). 65-75% of the percent of patients who do not respond do worse within first two years of treatment (Das et al,2024, ^4,5^). Therefore, identifying prognostic signatures to predict treatment response and disease recurrence and its applicability to Indian cohorts must be investigated if we were to tackle increasing incidence rates of breast cancer in India^6^. In a recent meta-analysis have shown that, compared to western cohorts, TNBC in Indian cohorts presents at an even earlier age (47 years vs 51 years), with high-grade tumors (2.57 OR) and greater lymph node involvement compared to non-TNBC patients ^7^. Most importantly, it was observed that TNBC prevalence is higher in India at 22-30% ^7–9^ as compared to 10-17% in western cohorts ^2^.

Previous studies in the western cohorts have investigated tumor heterogeneity at gene expression level to understand unpredictable treatment response and outcomes. Lehmann et al identified six subtypes of TNBC based on the gene-signature profiles ^10^, out of which four were tumor-specific subtypes; basal-like 1 (BL1), basal-like 2 (BL2) with high EGFR expression, mesenchymal (M), and luminal androgen receptor (LAR) with high AR expression, each showing distinct response to chemotherapy and survival outcomes ^11^. EGFR expression by immunohistochemistry in TNBC is higher ^12^ and is associated with poorer survival in TNBC cohorts from various ethnicities, including Egyptian, Greek, Chinese, and Japanese cohorts ^13–16^. Based on these studies, anti-EGFR and anti-AR trials have been initiated for TNBC subtype ^12,17–20^. There are currently at least three clinical trials investigating the efficacy of anti-EGFR therapies in TNBCs/basal breast cancers, all in phase 2. These trials have been focused on breast cancer patients in western cohorts. The expression of EGFR and AR in TNBC tumors in an Indian cohort and its effect on patient outcome is not well-studied in an Indian cohort.

In order to understand extent of AR and EGFR expression in TNBC from an Indian cohort, we set out to assess basal type (EGFR and/or CK5/6 positive) and LAR type (AR positive) TNBC in an Indian cohort of breast cancer patients. We further evaluated if these subtypes had implications towards patient outcomes. In a cohort of 93 TNBC, we noted CK5/6 expression had no correlation with survival outcomes. While EGFR-positivity was at 65% and AR-positivity was at 38%, both, with worse survival outcomes, though not significant. We also showed 25% of the TNBC tumors with clonal population of tissue and cell co-expression of EGFR and AR; such high frequency is unique to our cohort when compared to Western cohort data.

## Methods

### Sample selection

All the samples were taken from a biobank^21^ with appropriate ethics approvals (#IECHR/VB/2018/016 and an extension #EC/NEW/INST/2021/2443) built from a single surgeon’s practice in a tertiary breast cancer clinic. Formalin-fixed-paraffin-embedded blocks for invasive ductal carcinoma (IDC) and TNBC subtype were for patients diagnosed between 2012 to 19^th^ August 2022. Metastatic patients and samples of non-Indian origin were excluded from the cohort (Figure S1). All the samples were processed for this study after appropriate patient consent and study-specific ethics approvals.

### Collection of clinical, radiological, and follow-up data

Patient data such as age at diagnosis, menopausal status, tumor characteristics such as tumor grade, lymph vascular invasion (LVI) were obtained from patient clinical reports. Clinical tumor size and lymph node involvement data were obtained from sonomammography, mammography, and PET reports. Post-surgery pathological assessment showing overall tumor size and the number of involved lymph nodes to which the tumor had metastasized were also extracted for staging. Follow-up information of patients up to November 2023 was retrieved from the biobank database.

### Treatment regimens

Within this cohort, patients were treated by a single medical oncologist, therefore ensuring uniform treatment decisions according to NCCN guidelines for breast cancer ^22,23^. Despite the standard regimen recommendation, small variability in final treatment decision is observed due the socio-economic conditions of individual patients. In case of no lymph node involvement, treatment-naive tumor was surgically excised, followed by adjuvant chemotherapy (ACT) and radiotherapy where needed. In a cohort of 93 patients, 27 received NACT. AC (Anthracycline + Cyclophosphamide) + Taxane was the preferred chemotherapy option in both settings for 15 patients. Remaining 12 patients were given either AC alone, AC + 5-Fluoro-uracil, FEC (Fluoro-uracil + Epirubicin + Cyclophosphamide), or Gemcitabine.

### NACT treatment response

To compute the response for patients treated with NACT, we compared clinical and post-surgery pathological tumor size and node involvement status. Out of 27 NACT-treated patients, clinical (cTcN) and pathological stage (ypTypN) data was available for a subset 25 TNBC patients. On pathological assessment, if ypT0/ypTis and ypN0 and no metastasis was observed post-treatment, response was considered as pathological complete response (pCR). If downstaging of tumor was observed from cTcN it was recorded as partial response (PR). If no change in tumor size and number of nodes involved was observed, response was taken as stable disease (SD). In case of patients with increased tumor size and node involvement at post-surgery pathology report compared to what was reported at clinical diagnosis, response was reported as progressive disease (PD). Patients with partial response, stable, and progressive disease were pooled for final analysis into residual disease group since the number of patients in each group was too small to compare across variables.

### H&E and IHC staining and scoring

All the FFPE tissue were sectioned into 3-5 µm sections and H&E staining was done for all samples.

Immunohistochemistry for EGFR, CK5/6 and AR protein expression was standardized for each antibody (Table S1). DAB staining kit (Thermo Scientific, TL-125-QHD) was used to ensure uniform staining for all the samples.

Tumor and TILs percent for each sample and IHC expression scoring for each of the markers was done by a certified pathologist (AN).

### Whole-slide scanning

H&E and IHC slides were scanned at OptraScan facility using OptraSCAN, OS-15 bright field digital scanner at 400X resolution for whole-slide scanning. Images obtained at JP2000 (.jp2) are then converted to big tiff format.

### Multiplex Immunofluorescence staining (mIF)

To assess for co-expression of EGFR and AR protein within the tumor, tumor tissues with EGFR- and AR-positive IHC expression were stained for duplex EGFR and AR after optimization and validation with monoplex staining (Table S1). Duplex IF staining for AR and EGFR was done following the kit protocol from Akoya Biosciences, (OP7TL3001KT). Whole slide images of duplex staining were imaged with Leica Aperio VERSA automated scanning microscope. Images were processed on Aperio ImageScope software to identify areas of co-expression within the tumor tissue. Duplex images were annotated with tissue segmentation and cell segmentation based on DAPI staining, using HALO by Indica Lab version 3.6. EGFR-positive and AR-positive cells were quantified using the Highplex plug-in on HALO by Indica Lab version 3.6.

### FFPE RNA extraction, RNA-sequencing and analysis

Total RNA was extracted from 10-15µm curls from FFPE tissue samples. Out of 25 NACT-treated TNBC samples with response data available, RNA was isolated from eighteen samples for which tissue was available, using a standardized RNA extraction method. Quality assessment by RNA integrity number (RIN) and DV200 values was done using Agilent RNA 6000 Nano Kit on Agilent 2100 Bioanalyzer. cDNA library preparation was done using KAPA HyperPrep Kit for cDNA Synthesis & Amplification Module (KK8544) from 200 ng – 1 μg of total RNA. rRNA was removed by Ribodepletion using QIAseq FastSelect -rRNA (HMR) kit by suspending the input RNA in Fragment, Prime and Elute Buffer (1X). (Vaid et al, 2024, manuscript in submission). Upon passing the quality checks, all the samples were sequenced on Illumina platform by NovoSeq 6000. Raw reads were checked for base quality and adapter content using FastQC (v0.11.9). Fastp (v0.20.1) was used to remove adapter content and to trim low-quality bases. Hisat2 (v2.1.0) was used to map reads on the Homo Sapiens reference genome. Differential gene expression was performed using Deseq2 (v1.40.1) with a cut-off of FDR <0.1. Only samples which passed the sequencing quality check for higher mapping percentage and samples with higher gene count >5 were included in the final analysis.

### Co-*expression* analysis for EGFR and AR at cellular level using publicly available single-cell RNAseq

We downloaded the single-cell RNA sequencing raw data of TNBC patients from two different studies: (a) Wu et al. **[PMID 34493872]**^24^ and (b) Qian et al. **[PMID 32561858]**^25^. For Qian et al. dataset, count matrices from single-cell RNA sequencing (scRNA-seq) data of TNBC tumors (obtained using 10X v2 sequencing) were downloaded from http://blueprint.lambrechtslab.org along with cell annotations provided by the authors. To ensure data quality, the miQC package **[PMID 34428202]** was employed to eliminate non-viable cells, using a probability threshold of 0.5 to retain cells deemed to be of high quality. Likewise, for Wu et al. dataset, the count matrix and gene annotation were downloaded from the GEO ID GSE176078 (https://www.ncbi.nlm.nih.gov/geo/query/acc.cgi?acc=GSE176078).

Next, we created the Seurat Object using “Read10X” function for both the studies using Seurat version 4.0. As a part of quality control, we removed the cells which had more than 5% mitochondrial genes and the cells with unique feature count less than 200 or over 5000. To get TNBC specific samples with enrichment of cancer/epithelial cells we used ‘subset’ function on the selected samples. Next, we performed normalization and scaling to mitigate technical variability across cells. After Log normalization and scaling, principal component analysis (PCA) was performed. Subsequently, cluster of cells were characterized based on gene expression profiles using function ‘FindNeighbors’ with dims = 1:10 and ‘FindClusters’ with resolution as 0.5. Next, to visualize this high dimension data in lower-dimensional space to facilitate the exploration and interpretation of cell populations, we utilized dimensionality reduction method, uniform manifold approximation and projection (UMAP). ‘RunUMAP’ function was used to do the same with ‘reduction = pca’ and ‘dims = 1:10’. As our goal was to look at the cells expressing both Androgen Receptor (AR) and Epidermal Growth Factor Receptor (EGFR) genes, we extracted the list of ‘cell_ids’ expressing both the genes and highlighted them in the plot using “group.by” function. We also highlighted those cells which were expressing only AR or EGFR. Remaining cells were depicted as “Others” in the plot. “ggplot2” R package was used to create the final figures. The above analysis was performed for individual patient specific sample as well as for pooled samples in default manner (clusters of cells showing similar gene expression).

### EGFR and AR protein and RNA data from TCGA PanCancer dataset

To compare RNA and protein levels for EGFR and AR, z-scores from RPPA data and mRNA Expression levels from RSEM (Batch normalized from Illumina HiSeq_RNASeqV2) were downloaded from https://www.cbioportal.org/ (accessed on 15^th^ May 2024) for 874 breast cancer patients from TCGA BRCA-PanCan cohort ^26^. Associated clinical and histological information was obtained from GDC using TCGA bio links as described elsewhere ^27^. Only 429 IDC patients were included in the analysis for accurate comparison to our cohort. TNBC patients were identified if IHC showed ER-negative, PR-negative, and HER2-negative or her2 FISH-negative in case of samples with equivocal HER2 IHC scores. mRNA levels were log2 normalised before correlating with protein levels.

### Statistical analysis

All the statistical tests were done using GraphPad Prism v8.0. Chi-square test was done to assess unequal distribution of categorical variables. For continuous and discrete data, such as expression scores of EGFR, AR and CK5/6, percent tumor, Shapiro Wilk’s normality test was done to test if the data is normally distributed. If data was normally distributed, unpaired t-test was done to test mean differences between two variables and one-way ANOVA for more than two variables testing. If data was not normally distributed, Mann Whitney was performed to test the median differences between two variables and Kruskal Wallis to test median differences between more than two variables.

### Survival analysis

Disease outcomes were computed as follows: disease-free survival (DFS) was calculated as time in months from the date of surgery to the date of recurrence or last follow-up date. Recurrence within the first five years from the date of surgery is taken as an event. The patients who did not recur within the first five years from the date of surgery were censored at the of five years follow-up. Overall survival (OS) was calculated as time in months from the diagnosis date (biopsy date) till the last follow-up date or date of death (within the first five years). Kaplan-Meier survival plots for DFS and OS for up to 5-years follow-up time were plotted and Log-rank, Breslow, and Tarone-Ware computed survival probabilities towards 5-years DFS and OS. Log-rank Hazard ratio and 95% confidence interval (CI) were also computed for the higher hazard of having early DFS and OS. All plots were prepared using GraphPad Prism v.8.

## Results

### Triple-negative breast cancer cohort characteristics

EGFR, CK5/6 and AR expression was assessed by immunohistochemistry in a cohort of ninety-three TNBC patient samples from a breast cancer biobank ^21^. Demographic characteristics of the TNBC cohort are summarised in Table 1. At diagnosis, 45.45% of TNBC patients were below 50 years of age (n=48) and 40.79% were pre-menopausal (n=45). Three-fourths of TNBC patients had grade III primary tumors (76.09%, n=70). Upon assessment of clinical characteristics of the tumor, 69.5% of patients in the cohort presented with the larger tumor size, (cT2, n=57) and 71.08% had lymph node metastasis at the diagnosis (cN-positive, n=59). Therefore, higher number of patients in the cohort are presented with late stage (≥IIB) (63.75%) at diagnosis. Out of 93 patients, 27 patients underwent neo-adjuvant chemotherapy (NACT) followed by surgery, and 61 patients went for upfront surgery. Amongst the patients who went for upfront surgery, 68.72% patients had pathological tumor size between 2cm-5cm (pT2, n=33), 26.53% showed lymph node involvement (pN-positive, n=13) and 27.98% were late-stage patients (late pStage, n=13). Post-NACT, 42.31% (n=11) had ypT1 and 19.23% (n=5) were ypT2 stage, while 23.08% (n=6) patients had nodal involvement. 8% of NACT-treated patients were found to have pathological stage of IIB or above (n=2). The median follow-up time for the entire cohort is 29 months and the mean follow-up time is 35 months.

**Table 1:** Demographics of the cohort. Triple-negative breast cancer patient cohort characteristics are listed here. Patient data such as age, menopausal status, clinical parameters such as grade, radiological tumor size (cT), node status (cN) and stage, pathological parameters for NACT-treated (ypT, ypN, yStage) and untreated patients (pT, pN, pStage) for 93 TNBC patients is reported in categorical format. Proliferation marker – Ki67, Mesenchymal marker – Vimentin and Angiogenic marker – CD31 scores were obtained for all FFPE sections. Number of patients with positive and negative scores for each marker are listed here. Median and Mean follow-up for the 91 patients is shown here. Distribution across these parameters is shown as total number of patients (percent distribution). *NA includes patients with excision biopsy, n = 2

| **Total TNBC patients = 93** |  |  |
| --- | --- | --- |
| Clinical and pathological parameters |  | n (%) |
| Age, n = 88<br>(NA = 5) | Early age | 40 (45.45%) |
|  | Late age | 48 (54.55%) |
| Menstrual status, n=76<br>(NA=17) | pre-menopausal | 31 (40.79%) |
|  | post-menopausal | 45 (59.21%) |
| Grade, n=92<br>(NA=1) | Low | 22 (23.91%) |
|  | High | 70 (76.09%) |
| cT n=82<br>(NA=11) | cT1 | 19 (23.17%) |
|  | cT2 | 57 (69.51%) |
|  | cT3 | 5 (6.1%) |
|  | cT4 | 1 (1.22%) |
| cN n=83<br>(NA=10) | LN_negative | 24 (28.92%) |
|  | LN_positive | 59 (71.08%) |
| Clinical-Stage n=80<br>(NA=13) | Early stage | 29 (36.25%) |
|  | Late stage | 51 (63.75%) |
| NACT status n=88<br>(NA=5) | Yes | 27 (30.68%) |
|  | No | 61 (69.32%) |
| pT, n=48<br>(NA=18)* | pT1 | 12 (25%) |
|  | pT2 | 33 (68.75%) |
|  | pT3 | 2 (4.17%) |
|  | pT4 | 1 (2.08%) |
| pN, n=49<br>(NA=17)* | LN_negative | 36 (73.47%) |
|  | LN_positive | 13 (26.53%) |
| pStage, n=49<br>(NA=17)* | Early stage | 36 (72.92%) |
|  | Late stage | 13 (27.08%) |
| ypT, n=26<br>(NA=1) | ypT0 | 10 (38.46%) |
|  | ypT1 | 11 (42.31%) |
|  | ypT2 | 5 (19.23%) |
|  | ypT3 | 0 (0%) |
|  | ypT4 | 0 (0%) |
| ypN, n=26<br>(NA=1) | LN_negative | 20 (76.92%) |
|  | LN_positive | 6 (23.08%) |
| ypStage, NACT n=26<br>(NA=1) | Early stage | 24 (92.31%) |
|  | Late stage | 2 (7.69%) |
| Ki67, n=87 | Positive (>25%) | 64 (73.56%) |
| (NA=6) | Negative ($\leq 25\%$) | 23 (26.44%) |
| Vimentin, n=87<br>(NA=6) | Positive ($\geq 10\%$) | 47 (54.02%) |
| | Negative ($< 10\%$) | 40 (45.98%) |
| CD31, n=88<br>(NA=5) | Positive ($\geq 1\%$) | 28 (31.82%) |
| | Negative ($< 1\%$) | 60 (68.18%) |
| Follow-up period, n=91<br>(in months) | Median | 28.90 |
| | Mean ± S.D | 35.25 ± 30.87 |

Proliferation marker – Ki67, mesenchymal marker –Vimentin, and angiogenic marker -CD31, were also analysed by IHC on the serial sections of TNBC tumors within the same cohort. Ki67-positivity was noted for 64 (73.56%) patients, where more than 25% of the tumor section expressed Ki67(Figure S2A, Table 1). Vimentin-positivity was noted for 47 (54.02%) patients with 10% or more expression within tumor (Figure S2B, Table 1). For CD31, positivity was determined if any tumor cells expressed CD31 (≥1%), stromal CD31 was not considered as positive (Figure S2C, Table 1). 28 out of 93 TNBC patients were positive for CD31 (31.81%).

### EGFR-positive tumors were clinically aggressive and associated with worse survival

FFPE tumor sections of the TNBC patients from the cohort were stained for EGFR and CK5/6 expression by IHC to identify basal tumors. EGFR-positivity was determined when more than 10% of the tumor section had EGFR membrane expression (Figure 1A). Out of 93 TNBC samples, 65.2% of patients were EGFR positive for expression.

**Figure 1:**
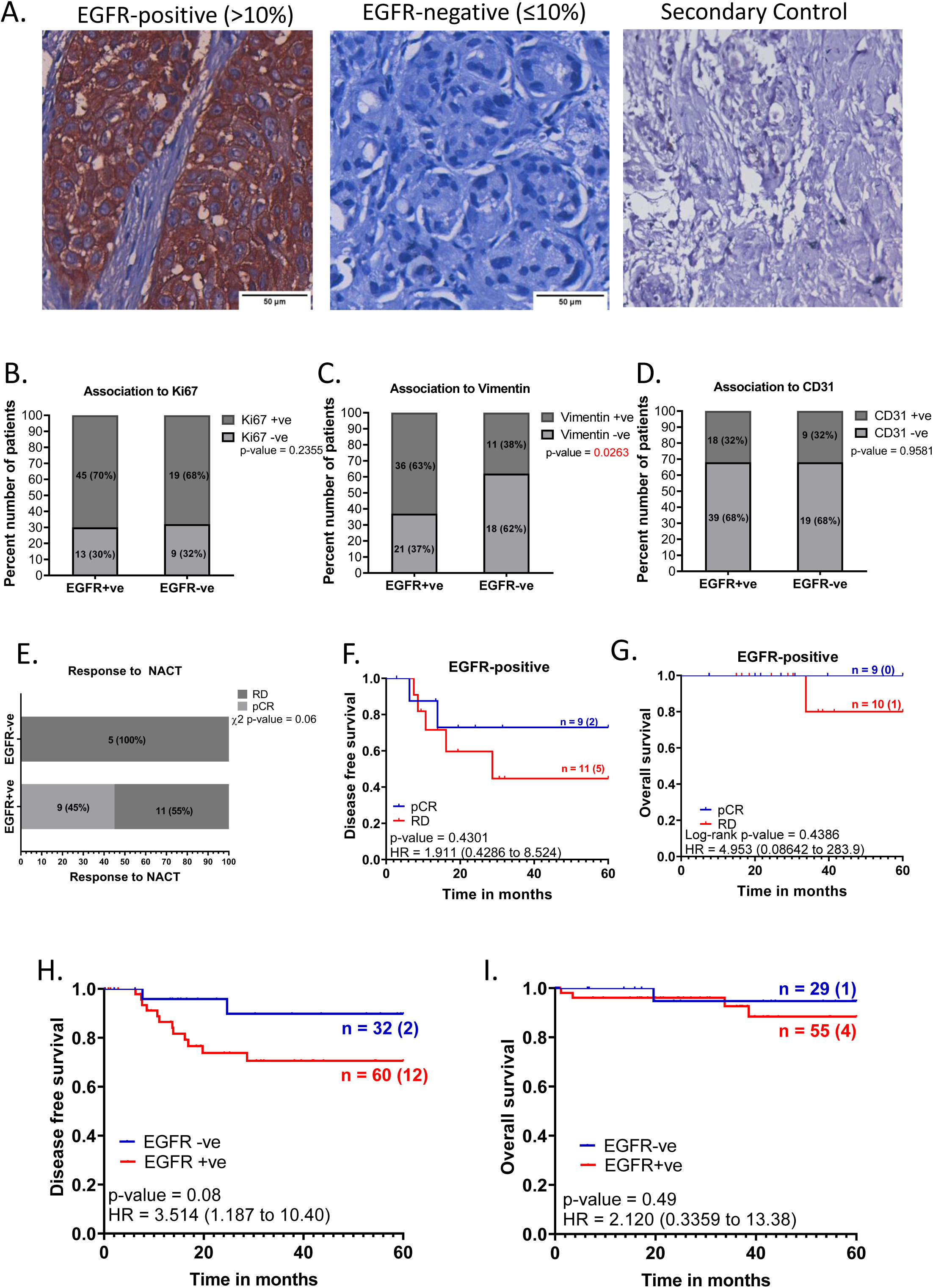
EGFR-positive TNBC patients were clinically aggressive and associated with poor survival. A. Representative images showing positive EGFR expression, negative EGFR expression and secondary control for IHC. Whole-slide scans of IHC slides were captured on OptraScan at 400X magnification and representative ROI are shown with the scale bar (50µm) added using Aperio ImageScope v12.4.3.5008. B, C, D: Distribution of EGFR +ve and -ve tumors across positive and negative scores of Ki67, Vimentin and CD31; respectively. v8. E: Response to NACT is shown here as pCR (pathological Complete response) and RD (residual disease) rate across EGFR+ve and EGFR-ve as horizontal stacked bars. For plots, B-E; p-values represent the χ^2^ test for the distribution of number of patients across the parameters compared here. In each bar, number of patients (percent number of patients) is shown. All graphs were prepared using GraphPad Prism. F-I: Survival outcomes plotted for EGFR+ve vs EGFR negative tumors: F. Disease-free survival and G. Overall survival outcomes across NACT-treated EGFR-positive tumors for patients who showed pCR or not (RD). H. DFS and I. OS across EGFR-positive and EGFR-negative tumors for the entire cohort of TNBC patients. Plots, log-rank p-value and Hazard ratio with respect to AR-negative tumors, along with confidence interval were analysed using GraphPad Prism 8.0.1.

To test if EGFR expression is associated with clinically aggressive tumors, EGFR-positive and EGFR-negative tumors were analysed for association across various clinical and pathological parameters (Table 2). We noted a trend, where EGFR-positive tumors were presented with a higher grade (50.55%, n=46) and late clinical-stage; <u>≥IIB</u> (43.04%, n=34) compared to EGFR-negative tumors

**Table 2:**
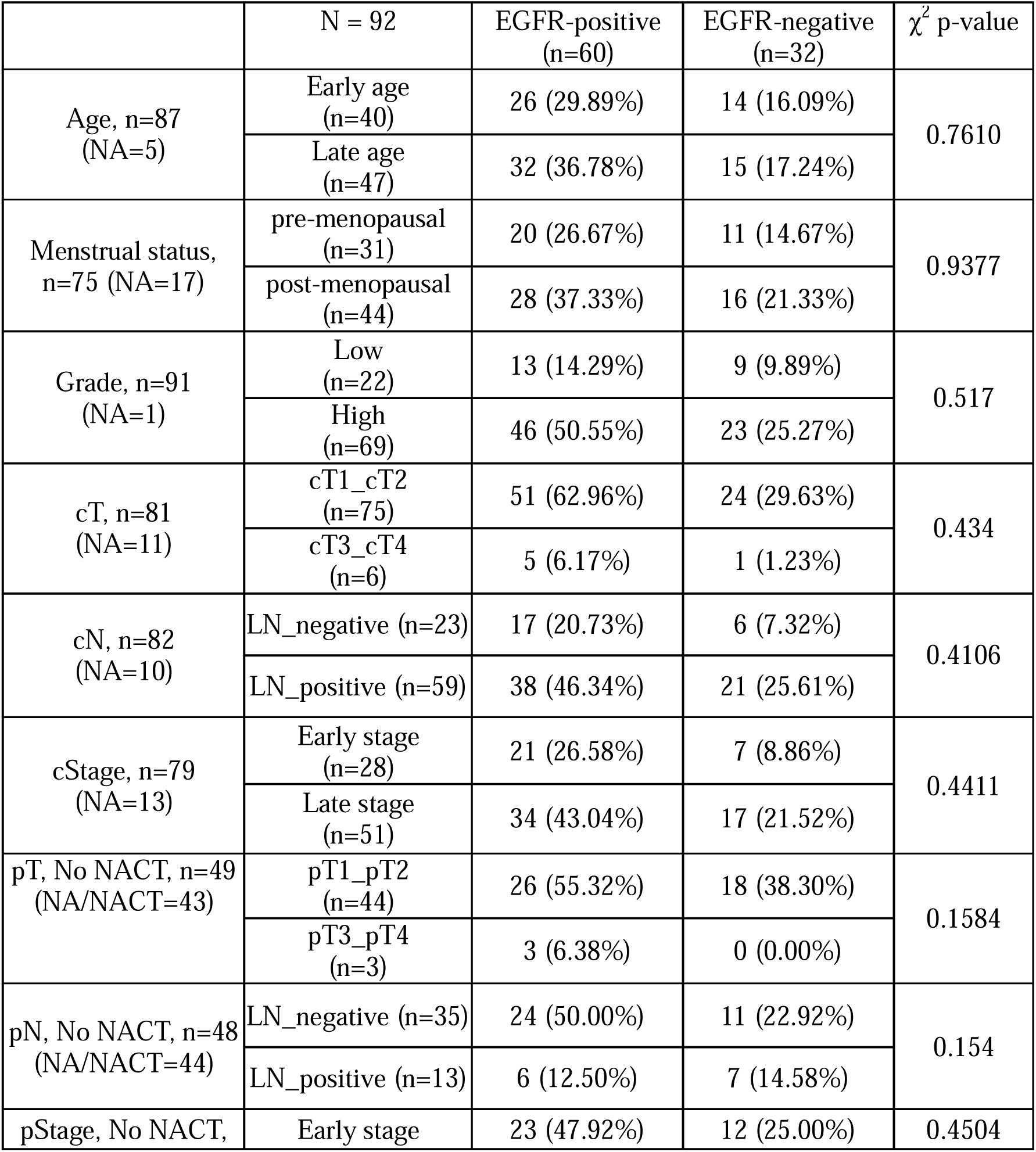
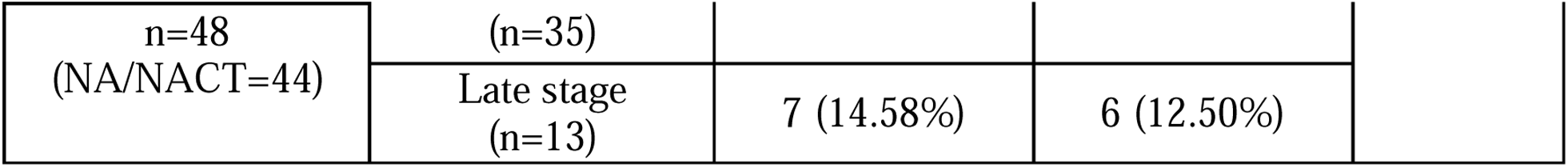
Clinical and pathological parameters distributed across EGFR-positive and EGFR-negative tumors. The number of patients across binned clinicopathological characteristics and percent distribution across EGFR+ and EGFR-tumors are noted in the table. Patient characteristics such as age, menopausal status and tumor grade at diagnosis, clinical features including tumor size (cT), lymph node involvement (cN) and clinical stage (cStage) and pathological features such as pT, pN and pStage were compared for categorical distribution using chi square test. χ^2^ test was computed using GraphPad Prism v8 for distribution across EGFR+ve and EGFR-ve tumors.

EGFR expression was analysed for association with treatment response for 25 patients treated with NACT. We observed that 45% (9 out of 20) of the EGFR-positive tumors showed complete response (pCR) compared to EGFR-negative tumors, where no patients showed pCR (0 out of 5) (p-value = 0.061) (Figure 1E). EGFR-positive patients with pCR show better disease-free and overall survival outcomes compared to EGFR-positive patients with residual tumors (RD) (Figure 1F and 1G).

For the overall cohort, we noted that EGFR-positive tumors showed around 20% higher recurrence rate compared to EGFR-negative tumors (p-value=0.078) with a hazard ratio of 3.514 (1.187 to 10.40) (Figure 1H). However, EGFR expression did not show any association with overall survival outcomes for the entire cohort of TNBC patients (Figure 1I).

Frequency of vimentin-positivity was significantly higher in EGFR-positive (63%, 36 out of 57, p-value = 0.026) tumors compared to that in EGFR-negative tumors (38%, 11 out of 29) (Figure 1C) indicating that EGFR tumors were more mesenchymal in nature. Ki67 and CD31 positive expression did not show any association with EGFR expression (Figure 1B, D).

Overall, in our cohort, EGFR-positive tumors were observed to be of mesenchymal nature and associated with poor disease-free survival.

### EGFR expression detects the clinical outcome of Basal tumors

Basal tumors were defined as with either EGFR (>10% membrane expression) and/or CK5/6 positive (>1%) expression. Non-basal tumors were defined with EGFR expression less than 10% and CK5/6 less than 1% (Figure 2A, B, C). 76 out of 88 tumors (81.7%) stained for either of the marker were identified as basal tumors, while 18.3% were negative for both the markers and were referred as non-basal tumors (Figure 2B and 2C).

**Figure 2:**
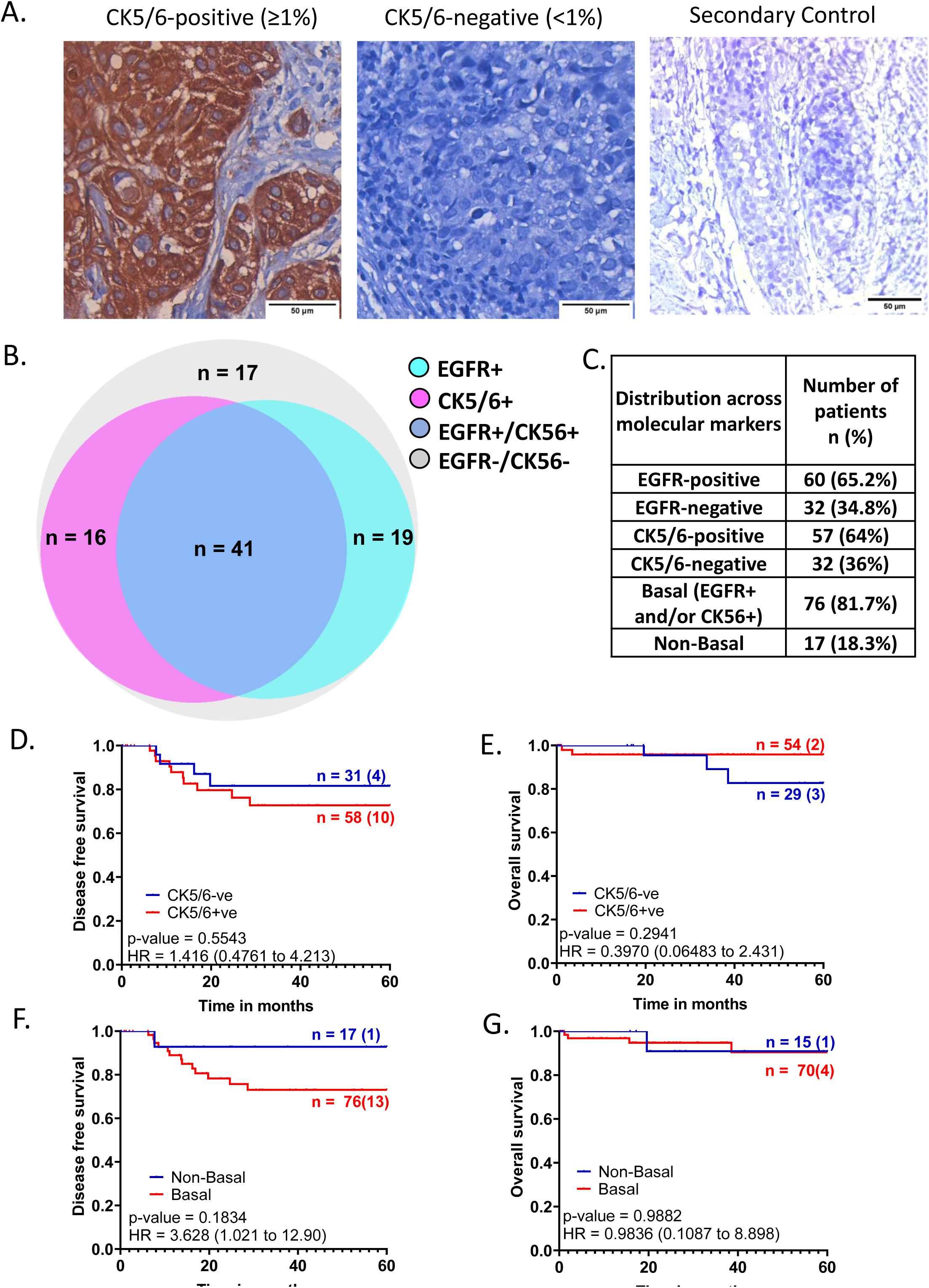
Basal tumor identification based on EGFR and CK5/6 marker expression and its distribution within the cohort. A. Representative images showing positive CK5/6 expression, negative CK5/6 expression and negative control. Whole-slide scans were captured on OptraScan at 400X magnification and representative ROIs are shown here with scale bar (50µm) added using Aperio ImageScope v12.4.3.5008. B. Venn diagram showing distribution of EGFR and CK5/6 positive tumors within the cohort. Figure made using <u>BioVenn program.</u> C. Table showing distribution of EGFR and CK5/6 positive tumors within the cohort and tumors with positive expression of both the markers. D., E.: KM plots depicting survival outcomes associated with CK5/6 expression and F., G.: Basal and non-tumors and non-basal tumors are plotted. The number of patients with event number in brackets is shown for each plot point. Plots, log-rank p-value and Hazard ratio, including confidence interval were analysed using GraphPad Prism 8.0.1.

Basal tumors were analysed for association with aggressive clinical characteristics (Table S2). Higher proportion of post-menopausal patients (47.37%, n=36) and higher-grade tumor were observed (64.13%, n=59) in basal tumors, similar to that of EGFR-positive tumors.

Basal tumors showed poor disease-free survival with a HR of 3.63 (CI - 1.021 – 12.90) for recurrence compared to non-basal tumors (Figure 2F). The long-term survival outcome for basal tumors associated with EGFR expression specifically as demonstrated by the outcome comparisons for EGFR or CK5/6 expression independently, as shown in Figures 1F and 2D. Unlike EGFR, CK5/6 positive tumors showed no association with the recurrence rate, with a hazard ratio of 1.41 (Figure 2D). No association was noted for Ki67 expression with basal and/or CK5/6 +ve tumors (Figure S3D and S3G). Basal tumors were significantly associated with high Vimentin expression scores compared to non-basal tumors (60%, n=43 vs 27%, n=4) (Figure S3H), suggesting they are more mesenchymal. We also noted that 64% of CK5/6 tumors (p-value = 0.0238) showed significant association with Vimentin-positive tumors (Figure S3E). A trend indicating a negative association between CD31 scores and basal tumors, compared to non-basal tumors (72%, n=52 vs. 50%, n=8) was observed (Figure S3I).

Overall, these results suggest that EGFR alone seems to be strongly associated with the patient survival outcomes within basal tumors, even though CK5/6+ tumors are more mesenchymal in nature. Therefore, in our cohort, EGFR alone stood out as an independent prognostic marker for TNBC patient survival outcomes.

### Androgen Receptor (AR) expression within TNBC tumors and patient outcomes

We assessed for the role of AR expression within TNBC cohort towards the clinical outcomes of TNBC patients. Of 93 samples stained for AR, 35 (38%) patients showed AR-positive expression with nuclear AR expression of more than ≥10% (Figure 3A). 46% of AR-positive tumors presented at early-stage, less than IIB, while 30% of AR-negative tumors presented at early stage (Table 3).

**Figure 3:**
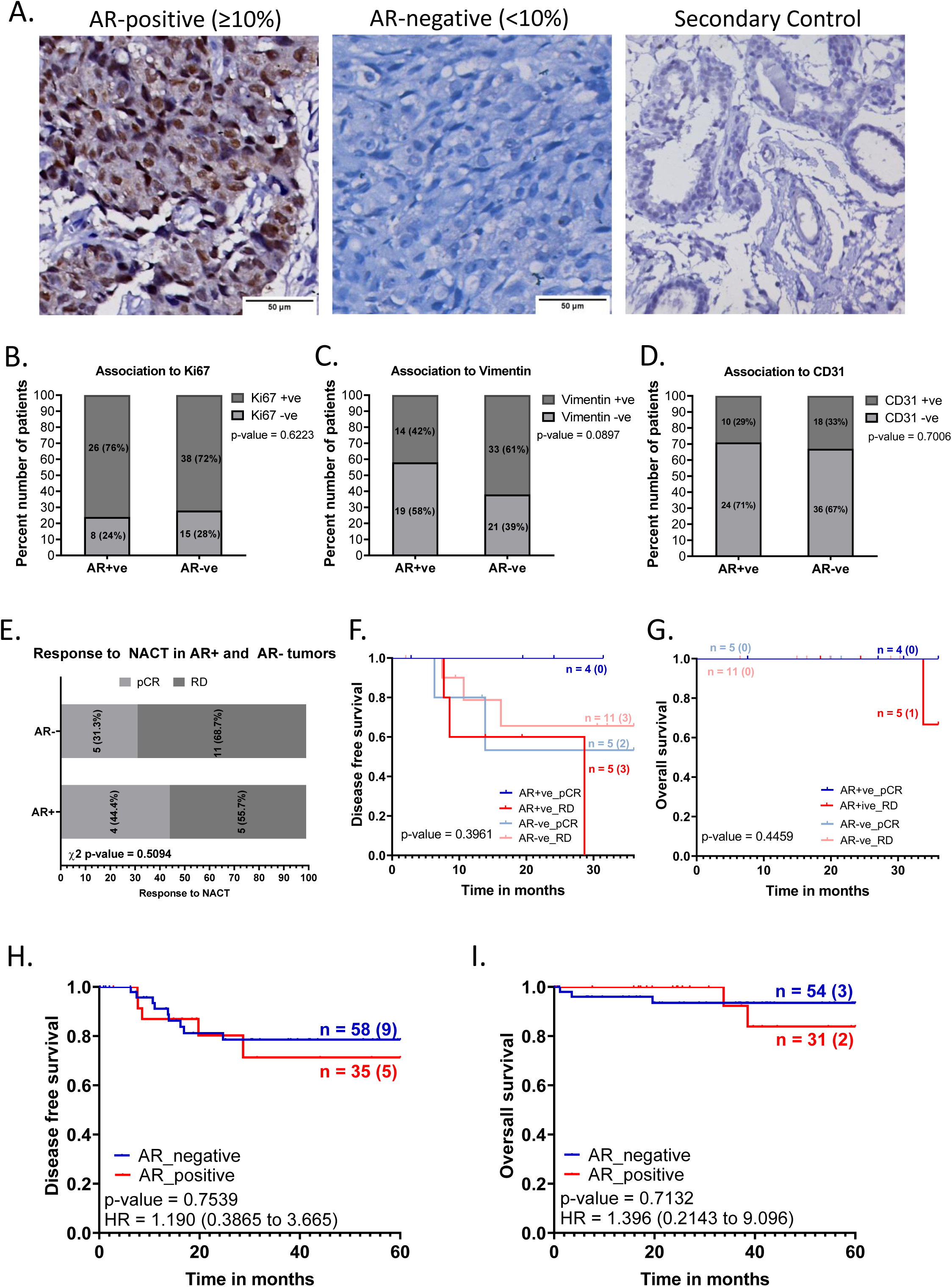
AR-positive TNBC patients were associated with poorer disease-free survival. A. Representative images showing positive AR expression, negative AR expression, and secondary IHC controls. Whole-slide scans were imaged using OptraScan at 400X magnification and representative ROI are shown with scale bar (50µm) added using Aperio ImageScope v12.4.3.5008. B, C, D: Distribution of Ki67, Vimentin and CD31 tumor expression across AR positive and negative tumors analysed with Chi square test. All graphs were prepared using GraphPad Prism v8. E: Response to NACT is shown here as rate of pCR (pathological complete response) and RD (residual disease) across AR+ve and AR-ve tumors. In each bar, number of patients (percent number of patients) is shown. F-I: Association of AR expression with survival outcomes: F. Disease-free survival and G. Overall survival outcomes across NACT-treated AR-positive tumors and AR-negative TNBC patients according to their pCR status. H. DFS and I. OS across AR-positive and AR-negative tumors for the entire cohort of TNBC patients. The number of patients with event number in brackets is shown for each plot point. Plots, log-rank p-value and Hazard ratio with respect to AR-negative tumors, along with confidence interval were analysed using GraphPad Prism 8.0.1.

**Table 3:** Clinical and pathological parameters distributed across AR-positive and AR-negative tumors. Table shows number of patients across clinicopathological characteristics and percent distribution across AR+ and AR-tumors. Patient characteristics such as age, menopausal status and tumor grade at diagnosis, clinical features including tumor size (cT), lymph node involvement (cN) and clinical stage (cStage) and pathological features such as pT, pN and pStage were compared here. χ^2^ test are computed using GraphPad Prism v8 to compare if any of the number of patients across the clinicopathological parameters compared here are unequally distributed between AR+ and AR-tumors.

| | N = 93 | AR-positive<br>(n=35) | AR-negative<br>(n=58) | $\chi^2$ p-value |
| --- | --- | --- | --- | --- |
| Age, n=88<br>(NA=5) | Early age<br>(n=40) | 12 (36.36%) | 28 (50.91%) | 0.2688 |
|  | Late age<br>(n=48) | 21 (63.64%) | 27 (49.09%) |  |
| Menstrual status, n=76<br>(NA=17) | pre-menopausal<br>(n=31) | 11 (40.74%) | 20 (40.82%) | >0.9999 |
|  | post-menopausal<br>(n=45) | 16 (59.26%) | 29 (59.18%) |  |
| Grade, n=92<br>(NA=1) | Low<br>(n=22) | 13 (38.24%) | 9 (15.52%) | >0.9999 |
|  | High<br>(n=70) | 21 (61.76%) | 49 (84.48%) |  |
| cT, n=84<br>(NA=11) | cT1-cT1<br>(n=76) | 28 (90.32%) | 48 (94.12%) | 0.6681 |
|  | cT3-cT4<br>(n=6) | 3 (9.68%) | 3 (5.88%) |  |
| cN, n=82<br>(NA=11) | LN_negative<br>(n=24) | 9 (28.12%) | 15 (30%) | >0.9999 |
|  | LN_positive<br>(n=58) | 23 (71.88%) | 35 (70%) |  |
| cStage, n=80<br>(NA=13) | Early stage<br>(n=29) | 14 (46.67%) | 15 (30%) | 0.155 |
|  | Late stage<br>(n=51) | 16 (53.33%) | 35 (70%) |  |
| pT, No NACT, n=49<br>(NA/NACT=40) | pT1-pT2<br>(n=45) | 15 (100%) | 30 (90.91%) | 0.5421 |
|  | pT3-pT4<br>(n=3) | 0 (0%) | 3 (9.09%) |  |
| pN, No NACT n=50 | LN_negative | 15 (100%) | 30 (90.91%) | 0.5421 |
| (NA/NACT=43) | (n=37) |  |  |  |
|  | LN_positive<br>(n=13) | 0 (0%) | 3 (9.09%) |  |
| pStage, n= 49<br>(NA/NACT=44) | Early stage<br>(n=36) | 13 (81.25%) | 23 (69.7%) | 0.5018 |
|  | Late stage<br>(n=13) | 3 (18.75%) | 10 (30.3%) |  |

Expression of Ki67, Vimentin, and CD31 were assessed for association with AR positivity within TNBC tumors. AR-positivity did not show a marked association to Ki67 expression in TNBC tumors. 76% of AR-positive tumors (26 out of 34) and 72% of AR-negative tumors (38 out of 53) were positive Ki-67 expression (Figure 3B). Vimentin was positive in 42% (14 out of 33 patients) AR-positive tumors compared to 61% (33 out of 54) of the AR-negative tumors. (Figure 3C). CD31 was low in both AR-positive (29%, 10 out of 34) and AR-negative tumors (33%, 18 out of 54), showing angiogenesis marker was independent of AR expression in tumors (Figure 3D).

For this cohort, we observed that AR-positive tumors were associated with marginally poorer disease-free and overall survival (Figure 3H and 3I). AR-positive tumors had lower disease-free survival probability of 0.78 (5 out of 35 recurred) compared to 0.71 for AR-negative tumors (9 out of 58 recurred) at 60 months of follow-up (Figure 3H).

Out of 27 patients who received NACT, 9 were AR-positive. Although this is a very small number to assess the response to treatment, we observed that the AR-positive patients showed a slightly higher rate of pCR (44.4%, n=4/9) compared to AR-negative tumors (31.3%, n=5/16) (Figure 3E). To understand whether there is any significant correlation between AR and NACT response, the effect of AR expression needs to be studied in a larger cohort of NACT-treated patients. Within AR-positive tumors, patients with residual disease (RD) had worse disease-free and overall survival compared to patients who achieved pCR (Figure 3F and 3G).

### Co-expression of EGFR and AR in TNBC tumors

Overall, our Indian TNBC cohort showed 38% AR-positivity and 64% EGFR-positivity. AR-positivity rate is noted to be higher than the previously reported studies ^28–31^ Lehman et al 2011 and 2016 classify TNBC tumors into distinct basal and LAR subtypes based on EGFR and AR expression, respectively, which precludes the possibility of co-expression of EGFR and AR. Interestingly, in our cohort, we observed a substantial fraction of tumors to co-express AR and EGFR. Twenty-five TNBC tumors out of ninety-three were positive (26.8%) for AR and EGFR expression on IHC assessment (representative images shown in Figure 4A).

**Figure 4:**
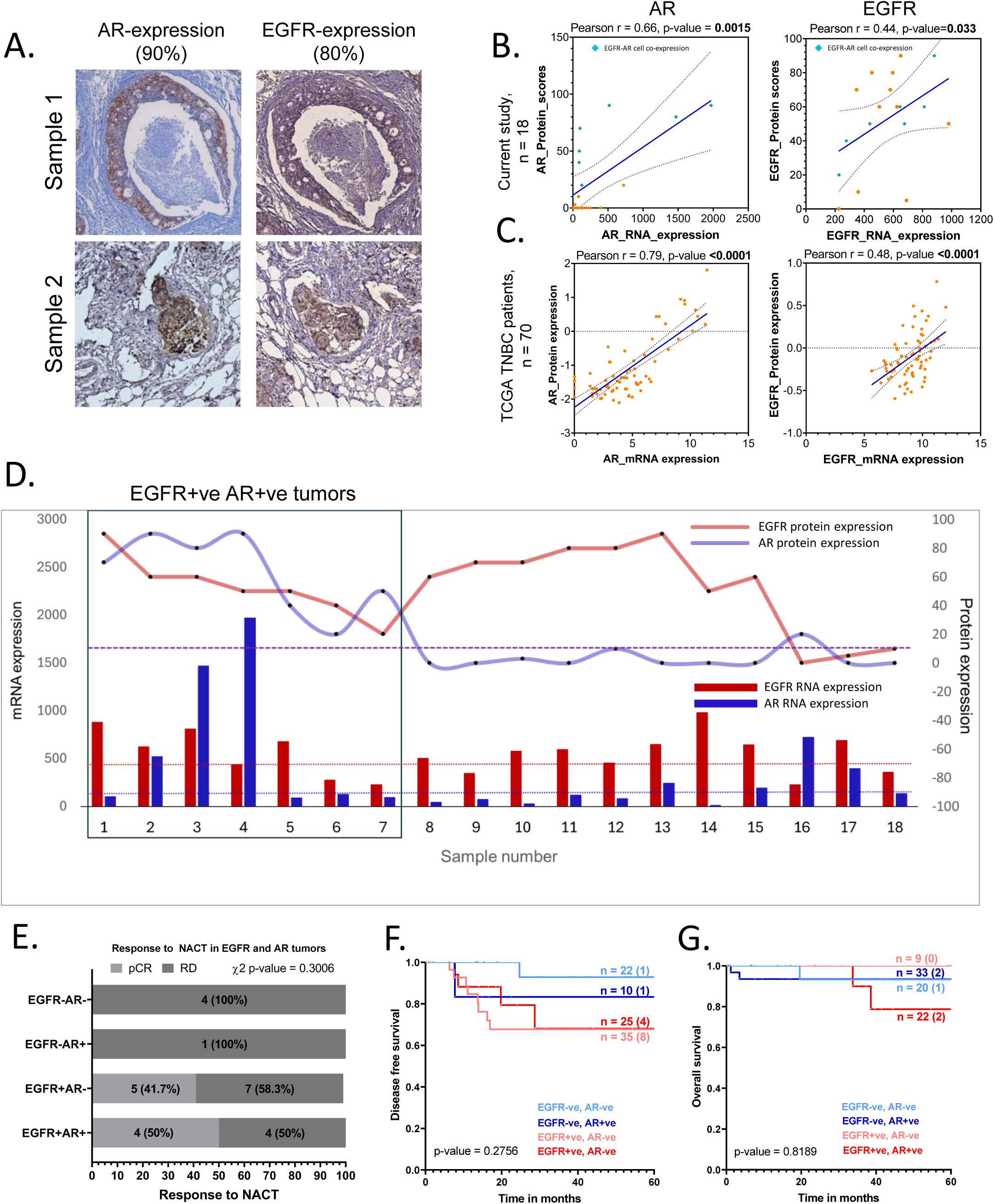
AR and EGFR tissue co-expressing TNBC tumors were clinically relevant. A. Representative IHC images taken at 40X showing AR and EGFR positive expression in the same ROI of adjacent serial sections of the same patient samples. AR and EGFR IHC scores are given on the top. Scale bar = 200µm. B. Scatter plots showing a linear correlation between protein and RNA expression levels of EGFR and AR in the study cohort. C. Correlation plots for protein expression levels detected by Reverse-phase protein assay (RPPA) and log2 (value+1) normalised mRNA expression levels for 70 TNBC patient samples from TCGA BRCA-IDC cohort. Orange dots show individual data points. Cyan dots show the data for samples with EGFR-AR cell-expression. The blue line indicates a linear regression line, and the dotted lines show confidence intervals. Pearson correlation tests were done to test if the two variables showed a significant correlation. R values and p-values are shown at the top left corner of the plots. Significant p-values are shown in bold. All graphs were prepared using GraphPad Prism v8. D. Plot showing EGFR and AR Transcript and Protein expression of individual patient sample. DE-seq2-normalized transcript levels are shown on Y-axis on the left, as bar graphs. Protein expression scores evaluated by IHC are shown on the Y-axis on the right, as black dots for individual samples joined by a smooth line. The red bar and line graph show EGFR RNA and protein levels. The blue bar and line graph show AR RNA and protein levels. Dotted red line indicates average EGFR transcript levels for all EGFR-ve samples and dotted blue line indicates average AR transcript levels for all AR-ve samples. The EGFR+ve AR+ve group is enclosed in a black box for representation purposes. E. Response to NACT is shown here as pCR (pathological Complete response) and RD (residual disease) rate across EGFR-AR-, EGFR-AR+, EGFR+AR- and EGFR+AR+ groups as horizontal stacked bars. F. Disease-free survival and G. Overall survival outcomes (right panels) across EGFR-AR-, EGFR-AR+, EGFR+AR- and EGFR+AR+ TNBC subgroups. The number of patients with event number in brackets is shown for each plot point. Plots and log-rank p-value along with confidence interval were analysed using GraphPad Prism 8.0.1.

Our current understanding is that TNBC that are of luminal origin express AR and those of basal origin express EGFR ^10,32,33^. In particular, the same tumor is not likely to express both the markers, unless it originates in both luminal and basal epithelium ^10,33^. To test if AR and EGFR transcript levels are indeed high in these tumors, we extracted total RNA from FFPE sections of these TNBC tumor blocks. Transcript levels were then compared with the protein expression by IHC for both the markers. We noted an RNA-to-protein concordance for individual TNBC samples as plotted in Figure 4D despite the heterogeneity. We did notice a significant concordance between EGFR and AR transcript and protein expression (Figure 4B). However, on closer inspection we found that residuals are non-randomly distributed (data not shown), and the relationship between transcript for AR transcript and protein is non-linear. The Spearman correlation between RNA and protein levels for AR was r = 0.66, and for EGFR was r = 0.40. We assessed the TCGA Breast Cancer dataset for their co-relation at mRNA and protein level, since the dataset had RNAseq and RPPA data available in the public domain for 429 IDC samples. (Figure 4C, Figure S4). We noted similar co-relation to our cohort. AR mRNA and protein levels were correlated within the entire IDC cohort with r = 0.76 (Figure S4A) and for the TNBC patients it was r = 0.78 (Figure 4B), while EGFR showed a correlation of r = 0.52 for the IDC cohort (Figure S4B) and r = 0.47 for TNBC patients (Figure 4B).

When assessed for NACT treatment response, EGFR and AR double-positive tumors showed marginally better pCR than EGFR-positive; AR-negative tumors (50% vs 42% pCR rate, respectively) (Figure 4E). Within AR+ve; EGFR-ve (n=1) and AR-ve; EGFR-ve (n=4) tumors, zero patients achieved pCR.

We also examined the association of co-expression of AR and EGFR-positive tumors for the entire TNBC cohort with disease-free and overall survival of the patients. (Figure 4F and Figure 4G). For Disease-free-survival (Figure 4F), EGFR-positive; AR-negative tumors showed worse outcomes for the first 2 years compared to that of EGFR-positive and AR-negative tumors, but this distinction was lost over the period of 5 years. While, EGFR-negative; AR-positive tumors did show worse DFS at the end of 5 years of follow-up time compared to that of EGFR-negative; AR-negative tumors (Figure 4F). For overall survival (Figure 4G), EGFR-positive; AR-positive tumors did worse compared to the rest.

### AR and EGFR cellular co-expression

To understand whether EGFR and AR co-expression was due to different cells of origin contributing to the clonal heterogeneity, we assessed the tumors for AR and EGFR expression using IHC and duplex staining. In 14 of the 25 tumors, we observed expression of AR and EGFR in concordant regions of the tumor in the serial sections (Figure S5). We validated this observation by performing duplex immunofluorescence staining for AR and EGFR on those 14 samples. Out of 14, tissue was available for further sectioning for 11 samples, which were processed for duplex staining. We obtained high-resolution whole slide scans for eight out of these 11 samples (Figure 5A and S6A).

**Figure 5:**
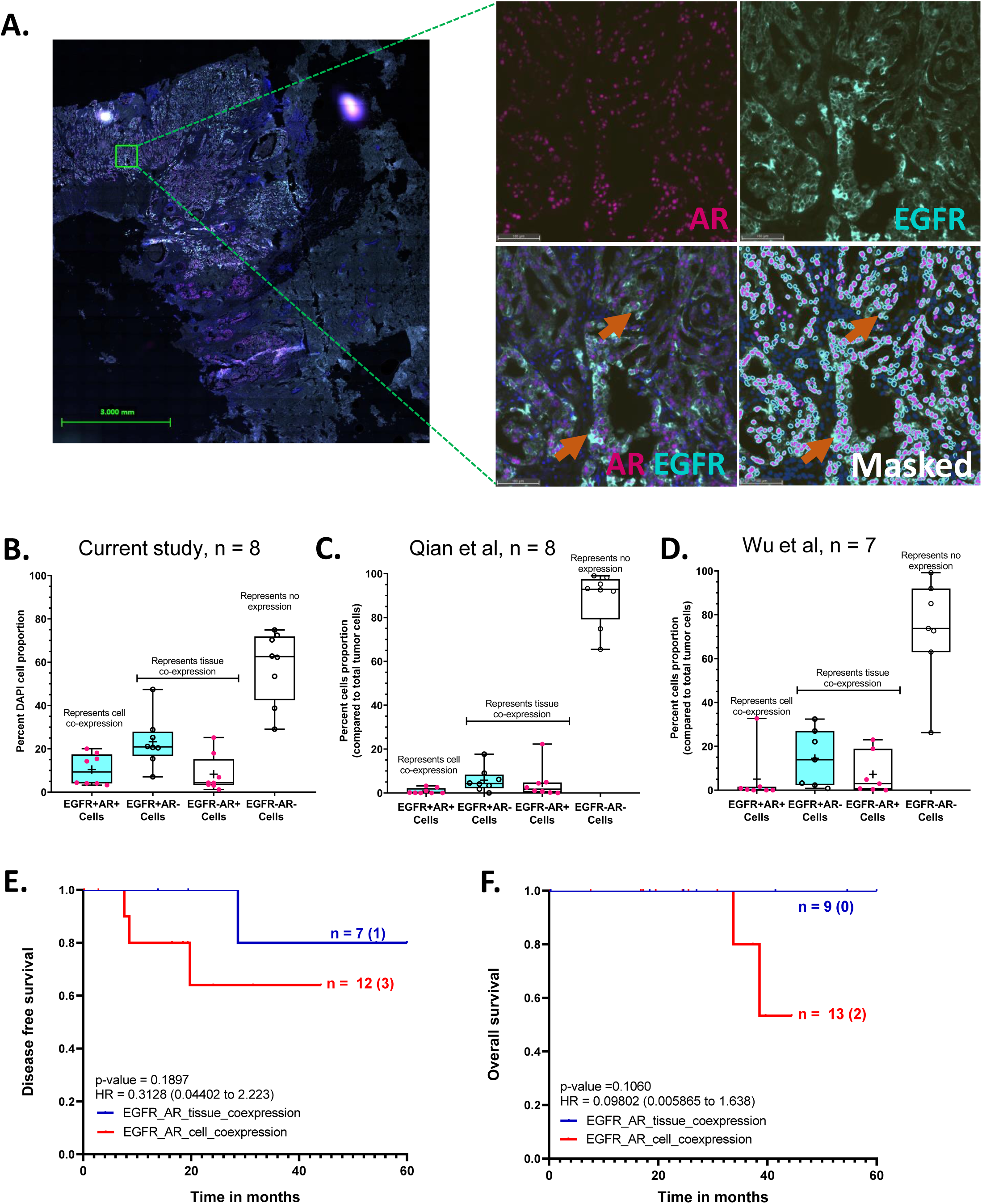
Hybrid cells co-expressing EGFR+AR+ cell were abundant and are associated with poor survival in TNBC patients. A. Representative image of multiplex IF-stained TNBC-tissue for AR and EGFR is shown here. Whole-slide scans were imaged using Leica Aperio VERSA 8 at 200X magnification (Scale bar – 3mm). Zoomed-in version of the inset image is shown on the right, was captured using Aperio ImageScope v12.4.3.5008. AR expression is shown in magenta and EGFR in cyan. Orange arrows indicate cells with AR and EGFR co-expressing tumor cells. All images were captured on HALO Indica software v3.6. Scale bar-100um. The figure at the right corner shows image with the mask for as quantified cells by Highplex plug-in on HALO for the respective marker. Box-whisker plots showing percent distribution of EGFR+AR+, EGFR+AR-, EGFR-AR+ and EGFR-AR-cell proportion for B. Eight TNBC samples within our cohort based on mIF expression where cell proportions were quantified on HALO Indica software v3.6 and total cell number were normalised to total DAPI+ve cell count; C. Eight TNBC samples from Qian et al study ^26^ based on scRNAseq expression normalised to total tumor cell count; D. Seven samples from Wu et al study ^25^ based on scRNAseq expression normalised to total tumor cell count. Cyan box indicates positive EGFR expression, white box indicates negative EGFR expression; pink dots indicate positive AR expression, white dots indicate negative AR expression. ‘+’ indicates mean values, and the median value is shown by the horizontal line in the box, while error bars indicate minimum and maximum cell proportion for all samples. E. Disease-free and F. Overall survival outcomes compared between patients with EGFR+AR tissue-(blue) and cell-(red) co-expression. Plots, log-rank p-value, Hazard ratio, and confidence interval were analyzed using GraphPad Prism 8.0.1.

We analysed the whole slide scanned images of EGFR and AR duplex staining with the Halo Indica software. We quantified the number of nuclei with EGFR-AR-, EGFR+AR-, EGFR+AR+ and EGFR-AR+ expression for each sample; we noted an average of 10.4% (range = 3.5 – 20.0%) of total DAPI+ cells with EGFR and AR co-expression, average 8.3% (range = 1.2 – 25.2%) of tumor cells with only AR expression and 23.3% (range = 7.1 – 47.4%) with only EGFR expression (Figure 5B). Around 58% (29.1 – 74.9%) of the remaining cell types (including non-tumor cell types such as stromal and immune cells) were negative for both EGFR and AR expression.

The exact origin and occurrence of such AR+EGFR+ cell population in TNBC tumors needs to be evaluated further. Within the scope of this study, we next aimed to assess if AR and EGFR co-expression are associated with patient prognosis and response to therapy.

Samples with AR+EGFR+ tissue co-expression and the ones with AR+EGFR+ cellular co-expression was analysed for survival outcomes. Samples with cellular co-expression had worse disease-free (p-value = 0.19) and overall survival (p-value = 0.11) compared to the patients with tissue co-expression (Figure 5E and 5F).

To assess if AR+EGFR+ cell population was present in other TNBC cohorts, we evaluated single-cell RNA sequencing datasets available for TNBC patients; Wu et al dataset ^30^ and Qian et al dataset ^31^(Figure S7). In Wu et al dataset, 4 out of 7 TNBC samples had cells co-expressing AR and EGFR (Figure S7B) with an average of 0.8% (range = 0 – 3.2%) cells showing co-expression (Figure 5C, Figure S7B). In Qian et al dataset, 5 out of 8 TNBC samples showed cellular co-expression (Figure 7C) with 5% (range = 0 – 32.7%) of total cells co-expressing EGFR and AR (Figure 5D, Figure S7D). In both the datasets, with the limited samples numbers, half of the TNBC patients exhibited cellular level co-expression of AR and EGFR genes (Figure S7). However, compared to the Indian cohort where 10.4% of cells showed co-expression, a small fraction of cells (0.8% and 5%) showed EGFR and AR co-expression in Wu et al and Qian et al study; respectively (Figures 5C and 5D).

Overall, we observed that high EGFR and AR expression in TNBC cohort, where EGFR positive expression was associated with higher recurrence rate. Serendipitously, we noted a proportion of TNBC patients co-expressing EGFR and AR within the same TNBC tissue with a portion of tumors showing cellular co-expression with a distinct association with the patient survival. We established our unexpected findings with the multiplex staining to confirm single-cell co-expression and validated it with the publicly available single-cell datasets. Therapeutic implications of such heterogenous cell population in TNBC, with co-expression of AR and EGFR needs to be explored further.

## Discussion

In this study, we profiled ninety-three TNBC patients from a cohort of breast cancer patients from India and classified them into basal/nonbasal and LAR subtypes based on of EGFR, CK5/6 and Androgen Receptor expression. We observed 65% EGFR-positive tumors and 81.7% were of basal origin within our cohort that expressed either EGFR or CK5/6, similar to previously reported for other Indian cohorts where EGFR-positive/ basal tumors ranged from 58 – 86% of the TNBC patients ^34–37^. This percent distribution is similar to what is reported to that of other non-Indian cohorts as well, where EGFR-positivity ranged from 60-71% ^13,16,38–40^. In concordance with these reports, we also noted poor association with disease-free survival, although the current cohort trend is statistically insignificant in our cohort.

Here we report the 38% AR-positive TNBC patients at 10% cut-off for AR-positive expression. To the best of our knowledge this is the highest proportion of AR positive cases in a TNBC cohort reported so far ^28–31,41–43^. In our cohort of 93 TNBC cases, the subset with high AR expression showed a less definitive association with the patient survival outcomes, although a trend towards shorter survival outcomes was observed. This is in contrast to AR associated outcomes reported for other TNBC cohorts, where AR expression was associated with better survival ^28,29,41^. Few exception to these reports are cohorts from India ^42,43^ and a study from US ^30^where survival outcome association with AR expression was not reported. The variable outcomes reported in these studies ^28–30,41^ are further validated by a study by Bhattarai et al in 2019^44^.

As per Lehmann et al.’s TNBC subtypes, LAR and Basal-like 2 - BL2 are two distinct TNBC subtypes with specific gene signatures where the LAR subtype is enriched for the AR gene expression, while high levels of EGFR expression were observed in the BL2 subtype^10^. In previous reports, basal TNBC has been identified based on EGFR and CK5/6 protein expression ^32,40^, while luminal TNBC is understood to express AR ^33^. While assessing EGFR and AR expression within TNBC tumors, we observed a unique subset of TNBC tumors with AR and EGFR co-expression at tissue and cellular levels. Furthermore, patients with cellular co-expression had worse survival outcomes than those with tissue co-expression.

We validated the cellular co-expression of EGFR and AR with multiplex immunofluorescence staining of tumor samples. To the best of our knowledge, tumor clones with double-positive EGFR and AR expression have not been reported elsewhere. We validated this novel observation in two independent single-cell RNAseq datasets available in the public domain ^24,25^ and confirmed the presence of similar cellular clones with EGFR and AR co-expression but to a lesser extent than our cohort. TNBC is well-known as a heterogenous tumor type ^45^. Tissue and cellular co-expression of EGFR and AR in 25% of the tumor samples from our cohort validates the complex heterogeneity within TNBC.

The presence of these clones, even in a small fraction (>1%), shows a higher hazard for recurrence in TNBC patients when compared to zpatients with tissue co-expression in our cohort. The exact biological implications of the EGFR+AR+ double positive tumor cells need to be further understood in in-vivo studies, which can help design better targeting strategies for such patients.

A few reports have assessed the expression of both markers in the same set of TNBC samples. In 2014, Thike et al reported AR expression in 39% of basal-like TNBC tumors at tissue level, with better survival outcomes compared to AR-negative basal tumors ^46^. Our cohort did not observe any difference in survival rates for AR-positive and AR-negative within basal/EGFR positive tumors. But in the absence of EGFR, patients with AR-positive tumors did marginally worse in our cohort, indicating that in the absence of EGFR, AR is involved in tumor progression and growth.

One major limitation of the current study is the small cohort size of TNBC patients. Although a trend is noted, we did not see a statistical significance towards survival for EGFR and AR or the double-positive tumor cells. Only 27 out of 93 patients were treated with neo-adjuvant chemotherapy within our cohort. Therefore, the association of EGFR or AR tumors to treatment response could be biased. A large-scale study with uniform distribution of NACT-treated patients with pCR and RD has to be conducted to validate our preliminary findings from the study.

Overall, we propose anti-AR trials should be taken seriously for breast cancer patients in India due to association with worse survival outcomes, as shown by others and our Indian cohorts studies. We propose, while designing anti-AR trials, double positivity of EGFR and AR should be considered to identify the group where targeting AR could be most effective.

This is one of the few molecular profiling studies for an Indian cohort of TNBC patients. We report 65 % of EGFR-positive and 38% AR-positive tumors in a cohort of 93 TNBC patients. Significantly, we identified a novel TNBC subgroup with EGFR and AR co-expression at tissue and at the cellular level with distinct association with survival outcomes.

## Data Availability

All data produced in the present study are available upon reasonable request to the authors.

http://blueprint.lambrechtslab.org

https://pubmed.ncbi.nlm.nih.gov/3https://www.ncbi.nlm.nih.gov/geo/query/acc.cgi?acc=GSE1760784493872/

https://doi.org/10.1016/j.cell.2018.02.052

## Acknowledgements

MK would like to acknowledge the DBT-Ramalingaswami ‘re-entry’ fellowship and DBT-Basic Research in Biology awarded by DBT-India. LSS would like to acknowledge DST-JC Bose Research Fellowship, Mphasis grant to TSB at Ashoka University. The research grant to CTCR is supported by Bajaj Auto Ltd. PV would like to acknowledge Ashoka University for PhD fellowship.

## Author contributions

Conceptualization: MK, PV, LSS, CK; Data curation: PK, SK, RB, RR, DK, MK; Formal analysis: MK, PV and PA; Funding acquisition: MK and LSS, CK; Investigation: PV, AK, AP, PA, MK; Methodology: PV, AK and MK; Project administration: MK and LSS; Resources: MK and LSS; Software: PV, PA; Supervision; MK and LSS; Validation: PA, PV, MK, DK; Visualization: PV, PA, DK, MK; Roles/Writing: original draft; PV, PA and MK; Writing - review & editing; PV, AP, PA, DK, SH, MK

## Data Accessibility

Publicly available single-cell data were obtained from the databases http://blueprint.lambrechtslab.org and <u>GSE176078</u>. RPPA data and mRNA Expression levels from RSEM (Batch normalized from Illumina HiSeq_RNASeqV2) were downloaded from https://www.cbioportal.org/). All additional data is included in the Supporting Information.

## Abbreviations

TNBC: Triple-negative breast cancer
EGFR: Epidermal Growth Factor Receptor
AR: Androgen receptor
IHC: Immunohistochemistry
DFS: Disease-free survival
OS: Overall survival
pCR: pathological Complete response

## References

1. Dent R, Trudeau M, Pritchard KI, et al. Triple-negative breast cancer: Clinical features and patterns of recurrence. Clin Cancer Res. 2007;13(15):4429–4434. doi:10.1158/1078-0432.CCR-06-3045

2. Foulkes WD, Smith IE, Reis-Filho JS. Triple-negative breast cancer. N Engl J Med. 2010;363(20):1938–1948. doi:10.1056/NEJMra1001389

3. Fournier M V., Goodwin EC, Chen J, Obenauer JC, Tannenbaum SH, Brufsky AM. A Predictor of Pathological Complete Response to Neoadjuvant Chemotherapy Stratifies Triple Negative Breast Cancer Patients with High Risk of Recurrence. Sci Rep. 2019;9(1):1–9. doi:10.1038/s41598-019-51335-1

4. Sharma RK, Gogia A, Deo S, Sharma D, Mathur S, Sagiraju HKR. Dose-dense neoadjuvant chemotherapy in triple-negative breast cancer: Real-world data from a developing country. Indian J Cancer. 2023;60(4):505–511. doi:10.4103/ijc.ijc_1120_21

5. Sharma S, Rathore SS, Verma V, Kalyan M, Singh N, Irshad I. Molecular Subtypes As Emerging Predictors of Clinicopathological Response to Neoadjuvant Chemotherapy (NACT) in Locally Advanced Breast Cancer (LABC): A Single-Centre Experience in Western India. Cureus. 2022;14(5):e25229. doi:10.7759/cureus.25229

6. Sung H, Ferlay J, Siegel RL, et al. Global cancer statistics 2020: GLOBOCAN estimates of incidence and mortality worldwide for 36 cancers in 185 countries. CA Cancer J Clin. Published online 2021. doi:10.3322/caac.21660

7. Kulkarni A, Kelkar DA, Parikh N, Shashidhara LS, Koppiker CB, Kulkarni M. Meta-Analysis of Prevalence of Triple-Negative Breast Cancer and Its Clinical Features at Incidence in Indian Patients With Breast Cancer. JCO Glob Oncol. 2020;(6):1052–1062. doi:10.1200/go.20.00054

8. Jonnada PK, Sushma C, Karyampudi M, Dharanikota A. Prevalence of Molecular Subtypes of Breast Cancer in India: a Systematic Review and Meta-analysis. Indian J Surg Oncol. 2021;12(April):152–163. doi:10.1007/s13193-020-01253-w

9. Suvobrata S, Murtaza A. Triple Negative Breast Cancer Prevalence in Indian Patients over a Decade: A Systematic Review. Int J Clin Biostat Biometrics. 2022;8(1). doi:10.23937/2469-5831/1510045

10. Lehmann BD, Bauer JA, Chen X, et al. Identification of human triple-negative breast cancer subtypes and preclinical models for selection of targeted therapies. J Clin Invest. 2011;121(7):2750–2767. doi:10.1172/JCI45014

11. Lehmann BD, Jovanović B, Chen X, et al. Refinement of triple-negative breast cancer molecular subtypes: Implications for neoadjuvant chemotherapy selection. PLoS One. 2016;11(6):1–22. doi:10.1371/journal.pone.0157368

12. Masuda H, Zhang D, Bartholomeusz C, Doihara H, Hortobagyi GN, Ueno NT. Role of epidermal growth factor receptor in breast cancer. Breast Cancer Res Treat. Published online 2012. doi:10.1007/s10549-012-2289-9

13. Abdelrahman AE, Rashed HE, Abdelgawad M, Abdelhamid MI. Prognostic impact of EGFR and cytokeratin 5/6 immunohistochemical expression in triple-negative breast cancer. Ann Diagn Pathol. 2017;28:43–53. doi:10.1016/j.anndiagpath.2017.01.009

14. Zhang L, Fang C, Xu X, Li A, Cai Q, Long X. Androgen receptor, EGFR, and BRCA1 as biomarkers in triple-negative breast cancer: A meta-analysis. Biomed Res Int. 2015;2015(Im). doi:10.1155/2015/357485

15. Liu D, He J, Yuan Z, et al. EGFR expression correlates with decreased disease-free survival in triple-negative breast cancer: A retrospective analysis based on a tissue microarray. Med Oncol. Published online 2012. doi:10.1007/s12032-011-9827-x

16. Levva S, Kotoula V, Kostopoulos I, et al. Prognostic evaluation of epidermal growth factor receptor (EGFR) genotype and phenotype parameters in triple-negative breast cancers. Cancer Genomics and Proteomics. 2017;14(3):181–195. doi:10.21873/cgp.20030

17. Yazdi MH, Faramarzi MA, Nikfar S, Abdollahi M. A comprehensive review of clinical trials on EGFR inhibitors such as cetuximab and panitumumab as monotherapy and in combination for treatment of metastatic colorectal cancer. Avicenna J Med Biotechnol. 2015;7(4):134–144.

18. Giusti RM, Shastri KA, Cohen MH, Keegan P, Pazdur R. FDA Drug Approval Summary: Panitumumab (Vectibix^TM^). Oncologist. Published online 2007. doi:10.1634/theoncologist.12-5-577

19. Kawaguchi Y, Kono K, Mimura K, Sugai H, Akaike H, Fujii H. Cetuximab induce antibody-dependent cellular cytotoxicity against EGFR-expressing esophageal squamous cell carcinoma. Int J Cancer. Published online 2007. doi:10.1002/ijc.22370

20. Wong SF. Cetuximab: An epidermal growth factor receptor monoclonal antibody for the treatment of colorectal cancer. Clin Ther. Published online 2005. doi:10.1016/j.clinthera.2005.06.003

21. Busheri L, Dixit S, Nare S, et al. Breast cancer biobank from a single institutional cohort in an urban setting in india: Tumor characteristics and survival outcomes. Cancer Treat Res Commun. 2021;28:100409. doi:10.1016/j.ctarc.2021.100409

22. National Comprehensive Cancer Network, Breast Cancer, Version 4.2017 — February 7, 2018. J R Soc Med. 2016;70(8):515–517. https://www2.tri-kobe.org/nccn/guideline/breast/english/breast.pdf

23. National Comprehensive Cancer Network. Breast Cancer, Version 32020. 2020;18(4):452–478. doi:10.6004/jnccn.2020.0016

24. Wu SZ, Al-Eryani G, Roden DL, et al. A single-cell and spatially resolved atlas of human breast cancers. Nat Genet. 2021;53(9):1334–1347. doi:10.1038/s41588-021-00911-1

25. Qian J, Olbrecht S, Boeckx B, et al. A pan-cancer blueprint of the heterogeneous tumor microenvironment revealed by single-cell profiling. Cell Res. 2020;30(9):745–762. doi:10.1038/s41422-020-0355-0

26. Liu J, Lichtenberg T, Hoadley KA, et al. An Integrated TCGA Pan-Cancer Clinical Data Resource to Drive High-Quality Survival Outcome Analytics. Cell. 2018;173(2):400–416.e11. doi:10.1016/j.cell.2018.02.052

27. Venkatasubramanian G, Kelkar DA, Mandal S, Jolly MK, Kulkarni M. Analysis of Yes-Associated Protein-1 (YAP1) Target Gene Signature to Predict Progressive Breast Cancer. J Clin Med. 2022;11(7). doi:10.3390/jcm11071947

28. Park S, Koo JS, Kim MS, et al. Androgen receptor expression is significantly associated with better outcomes in estrogen receptor-positive breast cancers. Ann Oncol. 2011;22(8):1755–1762. doi:10.1093/annonc/mdq678

29. Tang D, Xu S, Zhang Q, Zhao W. The expression and clinical significance of the androgen receptor and E-cadherin in triple-negative breast cancer. Med Oncol. 2012;29(2):526–533. doi:10.1007/s12032-011-9948-2

30. McGhan LJ, McCullough AE, Protheroe CA, et al. Androgen receptor-positive triple negative breast cancer: A unique breast cancer subtype. Ann Surg Oncol. 2014;21(2):361–367. doi:10.1245/s10434-013-3260-7

31. Pistelli M, Caramanti M, Biscotti T, et al. Androgen receptor expression in early triple-negative breast cancer: Clinical significance and prognostic associations. Cancers (Basel). 2014;6(3):1351–1362. doi:10.3390/cancers6031351

32. Cheang MCU, Voduc D, Bajdik C, et al. Basal-like breast cancer defined by five biomarkers has superior prognostic value than triple-negative phenotype. Clin Cancer Res. 2008;14(5):1368–1376. doi:10.1158/1078-0432.CCR-07-1658

33. Borri F, Granaglia A. Pathology of triple negative breast cancer. Semin Cancer Biol. 2021;72:136–145. 10.1016/j.semcancer.2020.06.005

34. Choccalingam C, Rao L, Rao S. Clinico-pathological characteristics of triple negative and non triple negative high grade breast carcinomas with and without basal marker (CK5/6 and EGFR) expression at a rural tertiary hospital in India. Breast Cancer Basic Clin Res. 2012;6(1):21–29. doi:10.4137/BCBCR.S8611

35. Rao C, Shetty J, Kishan Prasad HL. Immunohistochemical profile and morphology in triple - Negative breast cancers. J Clin Diagnostic Res. 2013;7(7):1361–1365. doi:10.7860/JCDR/2013/5823.3129

36. Sood N, Nigam JS. Correlation of CK5 and EGFR with clinicopathological profile of triple-negative breast cancer. Patholog Res Int. 2014;2014. doi:10.1155/2014/141864

37. Nag S, Dikshit R, Desai S, et al. Risk factors for the development of triple-negative breast cancer versus non-triple-negative breast cancer: a case–control study. Sci Rep. 2023;13(1):13551. doi:10.1038/s41598-023-40443-8

38. Park HS, Jang MH, Kim EJ, et al. High EGFR gene copy number predicts poor outcome in triple-negative breast cancer. Mod Pathol. 2014;27(9):1212–1222. doi:10.1038/modpathol.2013.251

39. Zhang M, Zhang X, Zhao S, et al. Prognostic value of survivin and EGFR protein expression in triple-negative breast cancer (TNBC) patients. Target Oncol. 2014;9(4):349–357. doi:10.1007/s11523-013-0300-y

40. Kanapathy Pillai SK, Tay A, Nair S, Leong CO. Triple-negative breast cancer is associated with EGFR, CK5/6 and c-KIT expression in Malaysian women. BMC Clin Pathol. 2012;12(1):1. doi:10.1186/1472-6890-12-18

41. He J, Peng R, Yuan Z, et al. Prognostic value of androgen receptor expression in operable triple-negative breast cancer: A retrospective analysis based on a tissue microarray. Med Oncol. 2012;29(2):406–410. doi:10.1007/s12032-011-9832-0

42. Saxena AK, Agarwal M, Singhal J, Gupta A. Evaluation of Androgen Receptor in Breast Carcinoma and its Correlations with Er, Pr, Her2/Neu, Triple Negative Receptor Status and Clinical Parameters. Int Arch Biomed Clin Res. 2017;3(3):48–51. doi:10.21276/iabcr.2017.3.3.13

43. Patnayak R, Jena A, Bhargavi D, Chowhan A. Androgen receptor expression in triple negative breast cancer - Study from a tertiary health care center in South India. Indian J Med Paediatr Oncol. 2018;39(1):28–31. doi:10.4103/ijmpo.ijmpo_4_17

44. Bhattarai S, Klimov S, Mittal K, et al. Prognostic role of androgen receptor in triple negative breast cancer: A multi-institutional study. Cancers (Basel). 2019;11(7):1–9. doi:10.3390/cancers11070995

45. Marra A, Trapani D, Viale G, Criscitiello C, Curigliano G. Practical classification of triple-negative breast cancer: intratumoral heterogeneity, mechanisms of drug resistance, and novel therapies. npj Breast Cancer. 2020;6(1):54. doi:10.1038/s41523-020-00197-2

46. Thike AA, Chong LYZ, Cheok PY, et al. Loss of androgen receptor expression predicts early recurrence in triple-negative and basal-like breast cancer. Mod Pathol. 2014;27(3):352–360. doi:10.1038/modpathol.2013.145

