## Supplementary file for "Molecular Profiling of a Triple-negative Breast Cancer Cohort in India for EGFR and AR expression analyzed for patient outcomes showed a distinct subset of cellular co-expression"

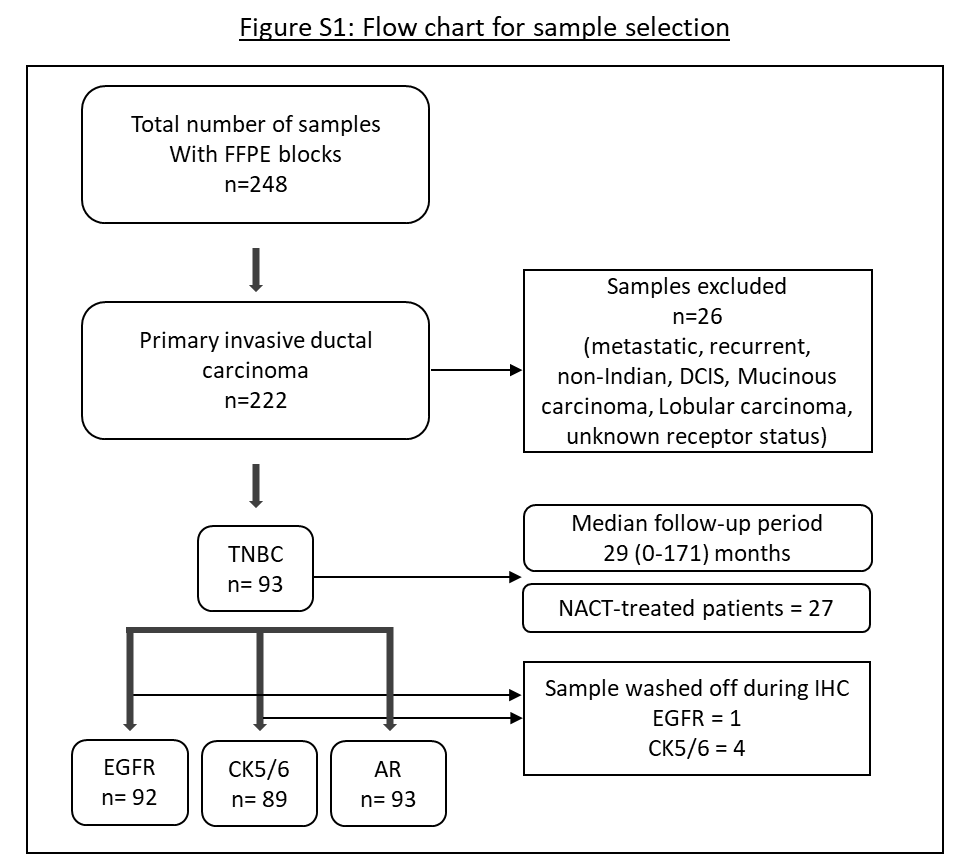

Figure S1: Flow chart for sample selection: Out of 248 FFPE block samples from PCCM biobank, 222 were IDCs, 93 of which were TNBC samples. Of all the TNBC samples stained for EGFR, CK5/6 and AR, one EGFR slide and four CK56 samples were washed off during staining.

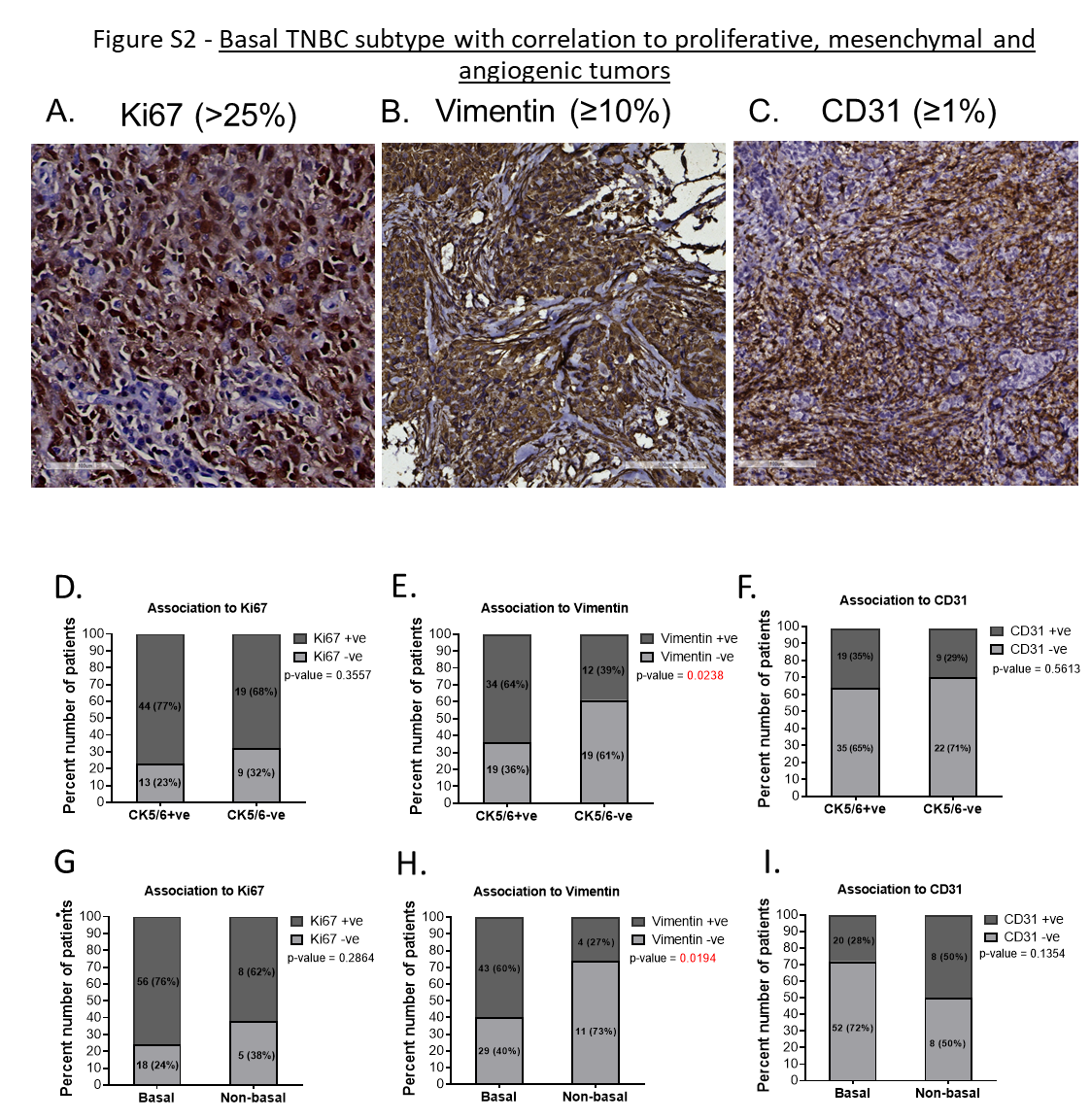

Figure S2: Basal TNBC subtype with correlation to proliferative, mesenchymal and angiogenic tumors.

A-B - Representative images showing positive expression of Ki67, Vimentin and CD31 for TNBC tumors. Whole-slide images were captured on OptraScan at 400X magnification, and representative ROI was selected, and a scale bar (100µm) was added using Aperio ImageScope v12.4.3.5008. D-I - Stacked bar graphs showing the distribution of Ki67+ve and -ve (left column), Vimentin+ve and -ve (middle column), and CD31+ve and -ve (right column) expression across CK5/6+ve and CK5/6-ve tumors (D-F) and across basal and non-basal tumors (G-I). p-value on the right top of all the graphs represents the χ2 value showing the distribution of number of patients across the parameters compared here. Red represents significant p-values. In each bar, number of patients (percent number of patients) is shown. All graphs were prepared using GraphPad Prism v8.

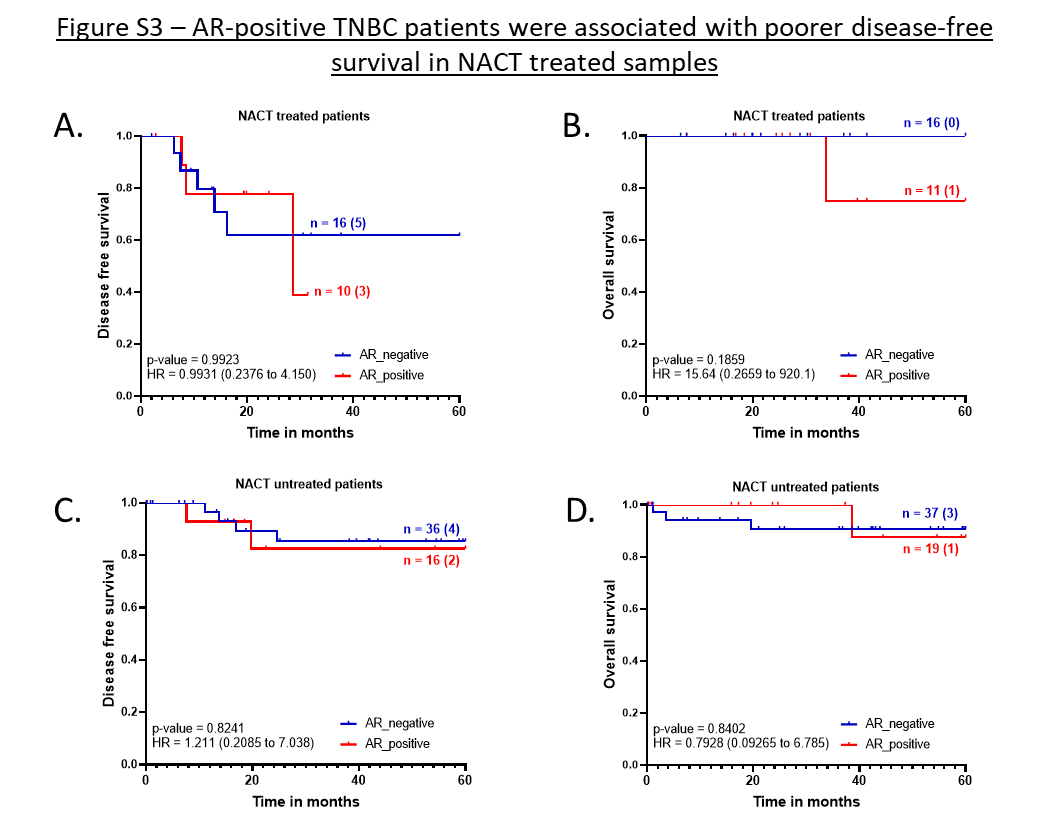

Figure S3: Survival outcomes of TNBC patients based on AR expression: Disease-free survival and Overall survival outcomes across A. B. NACT-treated AR-positive tumors and AR-negative TNBC patients according to their pCR status. A, C: DFS and B, D: OS across AR-positive and AR-negative tumors for the entire cohort of TNBC patients. The number of patients with event number in brackets is shown for each plot point. Plots, log-rank p-value and Hazard ratio with respect to AR-negative tumors, along with confidence interval were analysed using GraphPad Prism 8.0.1.

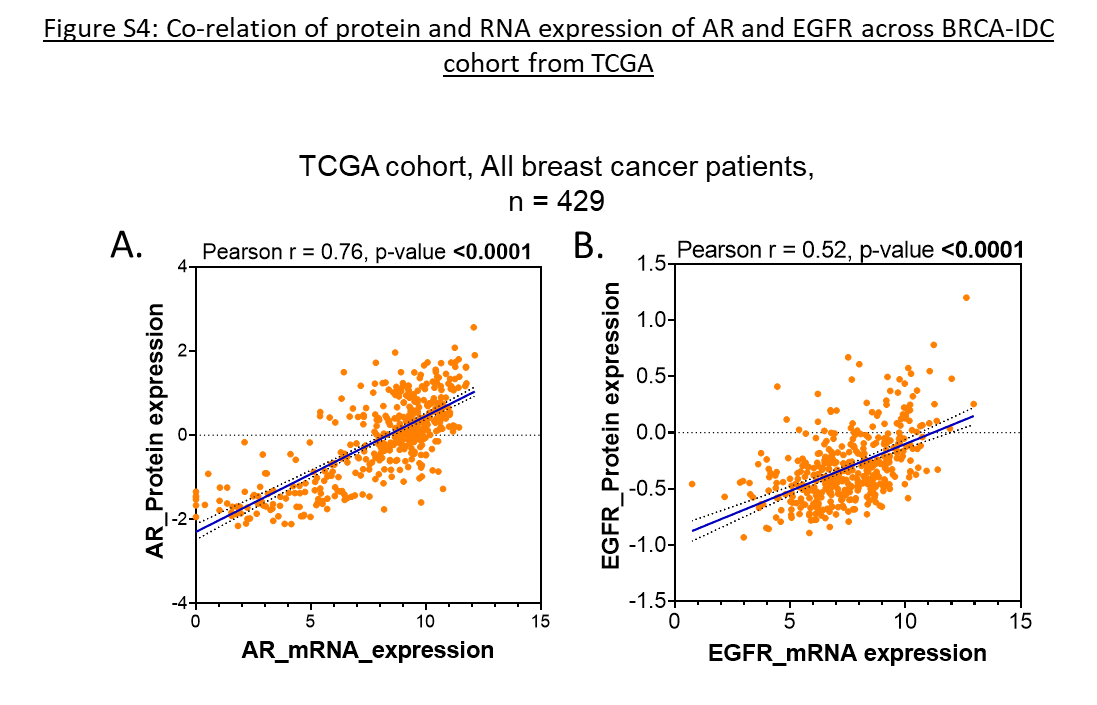

Figure S4: Co-relation of protein and RNA expression of AR and EGFR across BRCA-IDC cohort from TCGA. Scatter plots showing a linear correlation between protein and RNA expression levels for EGFR and AR. Correlation plots for A. AR and B. EGFR between protein expression levels detected by Reverse-phase protein assay (RPPA) and log2 (value+1) normalised mRNA expression levels for 429 patient samples from TCGA PanCancer cohort for all IDC breast cancer cohort. Orange dots shows individual data points. The blue line indicates a linear regression line, and dotted lines show confidence intervals. Spearman and Pearson correlation tests was done to test if the two variables showed significant correlation. R values and p-values are shown at the top left corner of the plots. Significant p-values are shown in bold. All graphs were prepared using GraphPad Prism v8.

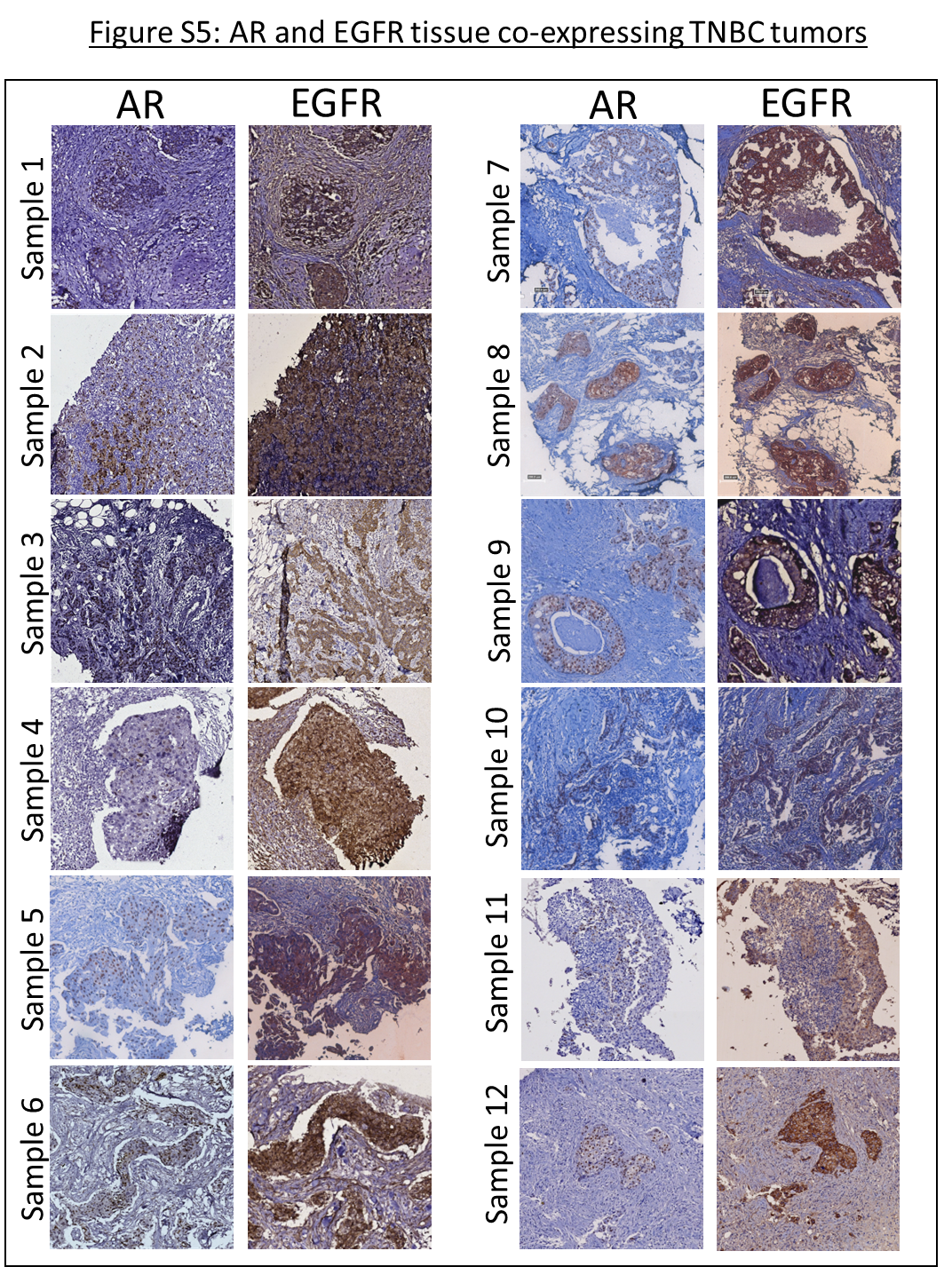

Figure S5: IHC expression of AR and EGFR in serial sections of TNBC tumors. Representative images showing AR+ and EGFR+ TNBC tumor sections stained with anti-AR and anti-EGFR antibody using DAB-based immune-histochemistry. Brown color indicates positive protein expression. Tumor cells showing co-expression of both the markers are comparable in the similar ROI in side-by-side panel for fourteen samples. Images were snapped from WSI images (captured by OptraScan at 400X magnification) at 10X optical zoom.

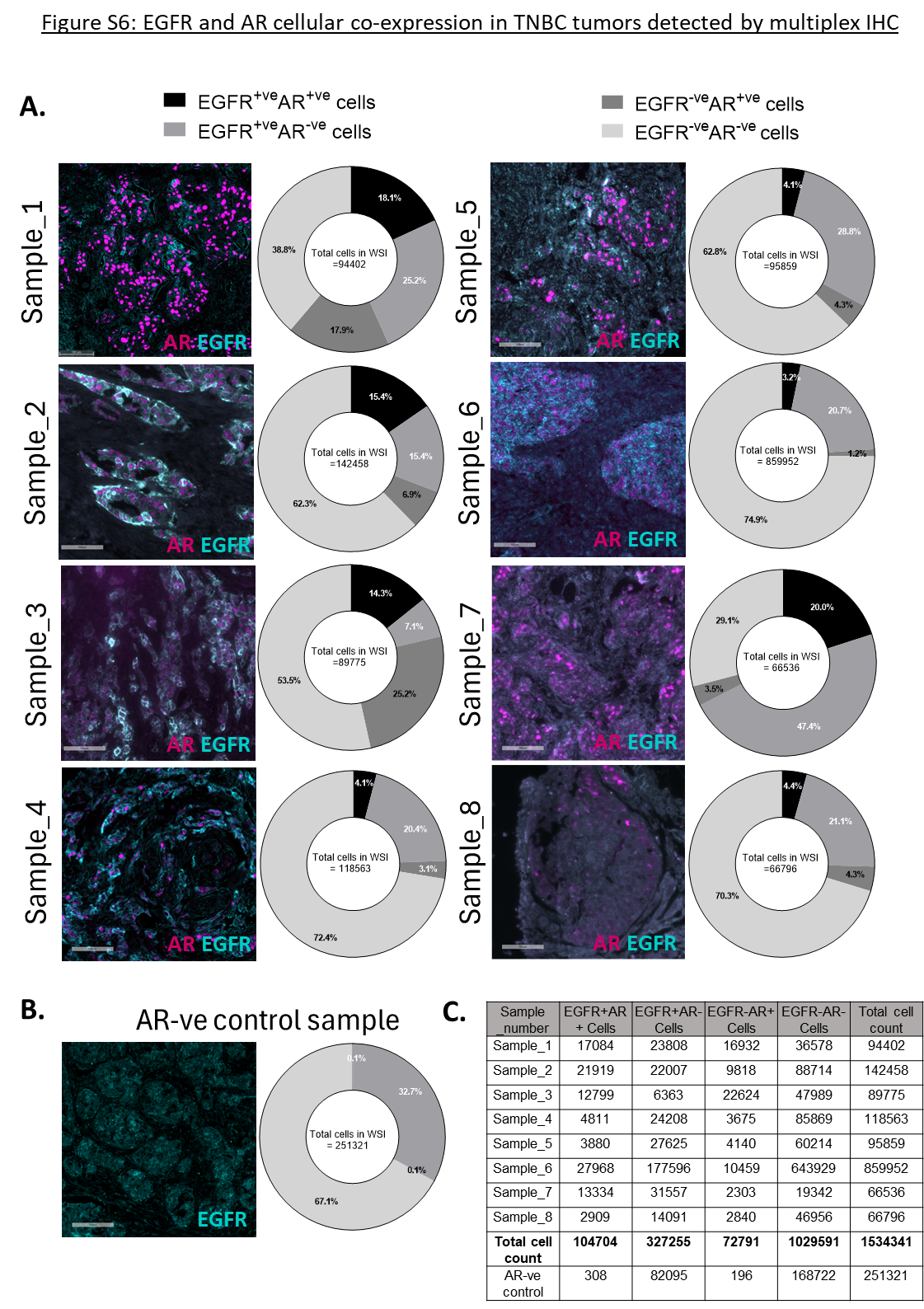

Figure S6: EGFR and AR cellular co-expression in TNBC tumors detected by multiplex IHC.

Representative ROI of EGFR and AR multiplex IF images of tumor sections for eight EGFR+AR+ TNBC samples (A) and one EGFR+AR-ve sample (B) are shown here. AR expression is shown in magenta and EGFR in cyan. Orange arrows indicate cells with AR and EGFR positive expression. WSI Images were captured on Leica Aperio VERSA 8. All images were processed on Aperio Imagescope v12.4.3. Scale bar-100um. Besides each image, donut graphs shows the percent distribution of EGFR+AR+, EGFR+AR-, EGFR-AR+, and EGFR-AR- for each sample as quantified by HALO Indica software v3.6. Total number of cells counted in each tissue image (total DAPI count) is shown in the centre. Donut graphs were plotted using GraphPad Prism 8.0.1. C. Raw cell count for each sample and total number of cells counted by multiplex imaging and analysis for the entire tissue for all the samples is shown in the table for reference.

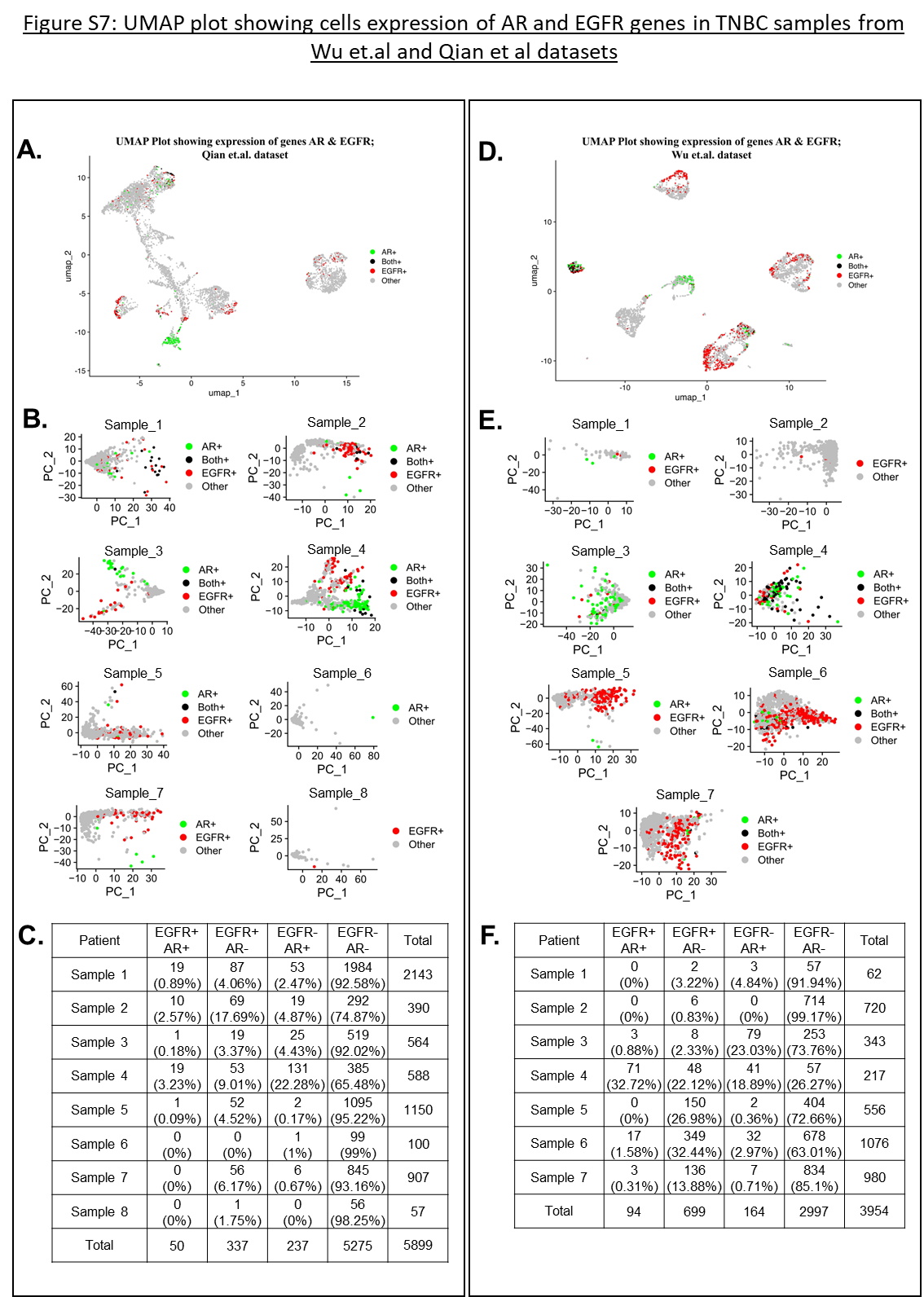
Figure S7: UMAP plot showing cells expression of AR and EGFR genes in TNBC samples from Wu et.al and Qian et al datasets.

A, B, D, E: UMAP showing the proportion of EGFR+, AR+, and EGFR+AR+ hybrid cells in external single-cell datasets for TNBC samples. The cluster formation is performed from A. B: Wu et. al dataset and D, E: Qian et.al dataset. A and D shows the clusters from compiled data from all the samples. B and E shows the clusters from individual patient samples. Cells expressing only AR gene are shown in green color; expressing EGFR in red color; both (AR & EGFR) in black color. Remaining cell types were denoted as “Others” and were shown in grey color. C and F: Table showing raw counts of total cells counted for each sample from both the datasets for EGFR+AR+, EGFR+AR- and EGFR-AR+ cells.

Table S1 - List of antibodies with staining conditions

| **Antibody** | **Company** | **Retrieval buffer** | **Antibody dilution** | **Primary antibody incubation time** |
| --- | --- | --- | --- | --- |
| AR, SP107 | ThermoFisher Scientific  (MA516412) | 1X Citrate Buffer (PathnSitu)  pH – 6 | 1:400 | 60 mins at RT |
| EGFR, EP38Y | Thermo Scientific  (RM-2111) | 10mM Tris-EDTA  pH – 9 | Ready-to-use | 20 min |
| CK 5/6, D5/16 B4 | Dako  (IS78030) | 10mM Tris-EDTA  pH – 9 | Ready-to-use | 60 mins at RT |
| Ki67, MIB-1 | Dako  (IS62630) | 10mM Tris-EDTA  pH – 9 | Ready-to-use | 60 mins at RT |
| Vimentin, V9 | Dako  (IS63030) | 10mM Tris-EDTA  pH – 9 | Ready-to-use | 60 mins at RT |
| CD31, JC70A | Dako  (IS61030) | 10mM Tris-EDTA  pH – 9 | Ready-to-use | 60 mins at RT |

| **Antibody for duplex IF staining** | **Company** | **Retrieval buffer** | **Antibody dilution** | **Primary antibody incubation time** |
| --- | --- | --- | --- | --- |
| AR, SP107 | ThermoFisher Scientific  (MA516412) | 1X Citrate Buffer (PathnSitu)  pH – 6 | 1:200 | 60 mins at RT |
| EGFR, EP22 | Master Diagnostica, MAD-000664QD-R-3) | 10mM Tris-EDTA  pH – 9 | 1:2 | 20 min |

Table S1 - List of antibodies with staining conditions: Table listing the antibody used and its clone number, company details, and catalog numbers. Retrieval buffer, pH, final dilution, and antibody incubation time are also stated for each antibody.

Table S2: Association of Basal and Non-basal tumor to clinicopathological characteristics

|  | N = 93 | Basal  (n=76) | Non-Basal  (n=17) | χ^2^ p-value |
| --- | --- | --- | --- | --- |
| Age,n=88  (NA=5) | Early age  (n=40) | 35 (39.77%) | 5 (5.68%) | 0.3006 |
|  | Late age  (n=48) | 38 (43.18%) | 10 (11.36%) |  |
| Menstrual status, n=78  (NA=17) | pre-menopausal  (n=31) | 26 (34.21%) | 5 (6.58%) | 0.6688 |
|  | post-menopausal  (n=47) | 36 (47.37%) | 9 (11.84%) |  |
| Grade,n=92  (NA=1) | Low  (n=22) | 16 (17.39%) | 6 (6.52%) | 0.2231 |
|  | High  (n=70) | 59 (64.13%) | 11 (11.96%) |  |
| cT,n=84  (NA=11) | cT1-cT2  (n=19) | 64 (92.75%) | 12 (80.00%) | 0.1272 |
|  | cT3-cT4  (n=57) | 5 (7.25%) | 3 (20.00%) |  |
| cN,n=83  (NA=10) | LN_negative  (n=24) | 18 (21.69%) | 6 (7.23%) | 0.3992 |
|  | LN_positive  (n=59) | 49 (59.04%) | 10 (12.05%) |  |
| cStage, n=80  (NA=13) | Early stage  (n=29) | 22 (27.50%) | 7 (8.75%) | 0.1493 |
|  | Late stage  (n=51) | 45 (56.25%) | 6 (7.50%) |  |
| pT, No NACT, n=49 (NA/NACT=40) | pT1-pT2  (n=12) | 35 (72.92%) | 10 (20.83%) | 0.3588 |
|  | pT3-pT4  (n=33) | 3 (6.25%) | 0 (0.00%) |  |
| pN, No NACT, n=48  (NA/NACT=40) | LN_negative  (n=35) | 29 (60.42%) | 6 (12.50%) | 0.3016 |
|  | LN_positive  (n=13) | 9 (18.75%) | 4 (8.33%) |  |
| pStage, No NACT, n=48 (NA/NACT=40) | Early stage  (n=35) | 29 (60.42%) | 6 (12.50%) | 0.3016 |
|  | Late stage  (n=13) | 9 (18.75%) | 4 (8.33%) |  |

Table S2: Association of Basal and Non-basal tumor to clinicopathological characteristics. Table show number of patients across binned clinicopathological characteristics and percent distribution across basal and non-basal tumors. Patient characteristics such as age, menopausal status and tumor grade at diagnosis, clinical features including tumor size (cT), lymph node involvement (cN) and clinical stage (cStage) and pathological features such as pT, pN and pStage were compared here. χ^2^ yest for done on GraphPad Prism v8 to compare if any of the parameters compared here are unequally distributed between basal and non-basal tumors.
